# Real-world effectiveness of sotrovimab for the treatment of SARS-CoV-2 infection during Omicron BA.2 and BA.5 subvariant predominance: a systematic literature review

**DOI:** 10.1101/2023.12.04.23299370

**Authors:** Myriam Drysdale, Mehmet Berktas, Daniel C. Gibbons, Catherine Rolland, Louis Lavoie, Emily J. Lloyd

**Affiliations:** Value Evidence and Outcomes, GSK, Middlesex, UK; Evidence Synthesis, Modelling and Communications, PPD Evidera, London, UK; Evidence Synthesis, Modelling and Communications, PPD Evidera, Montreal, Canada

**Author notes:** **Corresponding author:** Myriam Drysdale, GSK, 980 Great West Road, Brentford, Middlesex, TW8 9GS.

## Abstract

**Background:** Emerging SARS-CoV-2 variants have impacted the in vitro activity of sotrovimab, with variable fold changes in neutralization potency reported for Omicron BA.2 and subsequent variants. We performed a systematic literature review (SLR) to evaluate clinical outcomes associated with sotrovimab use during Omicron BA.2 and BA.5 predominance.

**Methods:** Electronic databases were searched for observational studies published in peer-reviewed journals, preprint articles and conference abstracts from January 1, 2022–February 27, 2023.

**Results:** The 14 studies identified were heterogeneous in terms of study design, population, endpoints and definitions, and comprised >1.7 million high-risk patients with COVID-19, of whom approximately 41,000 received sotrovimab (range n=20– 5979 during BA.2 and n=76–1383 during BA.5 predominance). Studies were from the US, UK, Italy, Denmark, France, Qatar, and Japan. Four studies compared the effectiveness of sotrovimab with untreated or no monoclonal antibody treatment controls, two compared sotrovimab with other treatments, and three single-arm studies compared outcomes during BA.2 and/or BA.5 versus BA.1. The remaining five studies descriptively reported rates of clinical outcomes in patients treated with sotrovimab. Rates of COVID-19-related hospitalization or mortality among sotrovimab-treated patients were consistently low (0.95% to 4.0% during BA.2; 0.5% to 2.0% during BA.5). All-cause hospitalization or mortality was also low (1.7% to 2.0% during BA.2; 3.4% during combined BA.2 and BA.5 periods). During BA.2, a lower risk of all-cause hospitalization or mortality was reported across studies with sotrovimab versus untreated cohorts. Compared with other treatments, sotrovimab was associated with a lower (molnupiravir) or similar (nirmatrelvir/ritonavir) risk of COVID-19-related hospitalization or mortality during BA.2 and BA.5. There was no significant difference in outcomes between the BA.1, BA.2 and BA.5 periods.

**Conclusions:** The studies included in this SLR suggest continued effectiveness of sotrovimab in preventing severe clinical outcomes during BA.2 and BA.5 predominance, both against an active/untreated comparator and compared with BA.1 predominance.

## Introduction

As of October 2023, there have been over 770 million confirmed cases of COVID-19 globally, including nearly 7 million deaths.^1^ Since the declaration of the COVID-19 pandemic by the World Health Organization (WHO) in March 2020,^2^ new severe acute respiratory syndrome coronavirus 2 (SARS-CoV-2) variants have continued to emerge.^3,4^ COVID-19 continues to be responsible for a substantial number of new infections globally, placing a strain on healthcare systems around the world.^1,5^

Sotrovimab is a dual-action recombinant human IgG1κ monoclonal antibody (mAb) derived from the parental mAb S309, a potent neutralizing mAb directed against the spike protein of SARS-CoV-2.^6–9^ The safety and efficacy of sotrovimab was demonstrated in the pivotal COMET-ICE randomized clinical trial (NCT04545060), conducted during the original ‘wild-type’ variant period of the pandemic.^10^ A single intravenous (IV) infusion of sotrovimab (500 mg) was found to significantly reduce the risk of all-cause >24-hour hospitalization or death by 79% compared with placebo in a high-risk population with COVID-19.^10^ Sotrovimab (IV 500 mg) was subsequently granted Emergency Use Authorization (EUA) by the United States (US) Food and Drug Administration (FDA) for the treatment of mild-to-moderate COVID-19 in adults and pediatric patients (≥12 years of age and ≥40 kg) who tested positive for SARS-CoV-2 and were at a high risk of progression to severe COVID-19, including hospitalization or death.^11^ Sotrovimab was also granted marketing authorization in the European Union, Norway and Iceland,^12^ and Bahrain, and conditional marketing authorization in Australia,^13^ the United Kingdom,^14^ Saudi Arabia and Switzerland.^15^ In Japan, a Special Approval in Emergency has been granted, and temporary/emergency authorizations were granted in Canada, and the United Arab Emirates.

Since the COMET-ICE trial was undertaken, new viral variants have emerged, including the Omicron BA.2 subvariant that became predominant globally in March 2022^16^ and the BA.5 subvariant that became predominant in August 2022.^17^ In vitro neutralization assays demonstrated that sotrovimab retained its neutralization capacity against Omicron BA.1 but showed reduced neutralization potency against later variants, such as Omicron BA.2 and BA.5 (16- and 22.6-fold changes in EC50, respectively).^18^ In the absence of clinical trials to assess the efficacy of sotrovimab against these emerging variants, the clinical relevance of this reduced neutralization observed in vitro was unknown, and the FDA took the decision in April 2022 to deauthorize the EUA for sotrovimab in the US.^19^

Generating near real-time data on the efficacy of sotrovimab in the constantly evolving SARS-CoV-2 variant landscape is challenging, and there is no validated model that can reliably correlate in vitro neutralization to predicted clinical efficacy; hence, real-world evidence is a key source of information to assess the benefit-risk profile of sotrovimab. A published systematic literature review (SLR) and meta-analysis of 17 studies including 27,429 patients concluded that sotrovimab is an effective and well-tolerated therapy that can reduce mortality and hospitalization rates in patients infected with both the Delta and Omicron BA.1 variants.^20^ In addition, we previously conducted a SLR of papers published from January 1^st^ to November 3^rd^, 2022, the results of which suggested continued clinical effectiveness of sotrovimab in preventing severe clinical outcomes related to COVID-19 during Omicron BA.2 predominance versus a control/comparator and compared with the period of BA.1 predominance.^21^

To investigate the use of sotrovimab against emerging variants among patients either partially or fully vaccinated against or previously exposed to SARS-CoV-2, including impact on clinical outcomes, a SLR was undertaken to evaluate the current evidence on the clinical effectiveness of sotrovimab during Omicron BA.2 and BA.5 predominance. This SLR builds on our previous review^21^ to cover studies including BA.5 predominance periods and newly published papers on BA.2.

## Methods

This SLR included observational studies investigating clinical outcomes in patients treated with sotrovimab published in peer-reviewed journal articles, preprint articles, and conference abstracts between January 1, 2022 and February 27, 2023. The publication period was selected to identify publications reporting data during Omicron BA.2 and BA.5 predominance. Where available, data on other circulating variants were also extracted for potential comparison between periods of variant predominance.

The SLR was conducted in accordance with Preferred Reporting Items for Systematic Reviews and Meta-Analyses (PRISMA) guidelines (PROSPERO registration number: CRD42022376733).^22^

### SLR objectives

The primary objective of the SLR was to assess the clinical effectiveness of sotrovimab in patients receiving early treatment for COVID-19 (as used in accordance with local COVID-19 guidelines) during the Omicron BA.2 and BA.5 predominance periods.

### Data sources and search strategy

Searches were conducted using the following indexed electronic databases: MEDLINE (via OVID), Embase (via OVID), LitCovid (via MEDLINE), Cochrane COVID-19 Study Register, and EconLit. Additional searches for relevant preprints were conducted in ArRvix, BioRxiv (via Embase), ChemRvix, MedRxiv (via Embase), Preprints.org, ResearchSquare, and SSRN.

The following conferences were also searched for relevant abstracts indexed from January 1, 2022: Infectious Diseases Week; International Conference on Emerging Infectious Diseases; European Respiratory Society; and European Congress of Clinical Microbiology and Infectious Diseases. These conferences were selected as they were likely to include a wide range of newly available research in the field of COVID-19 therapeutics and management.

Search strategies, starting from January 1, 2022 for each database, included a combination of free-text search terms for COVID-19, different variants, sotrovimab, and observational study design (**Supplementary Table 1**). There was no limit on geographical location, but only English language publications were considered.

### Study selection

Studies were screened and selected for inclusion in the SLR against predetermined PICOS (populations, interventions and comparators, outcomes, and study design) criteria.^23^ Only studies matching any inclusion criteria and none of the exclusion criteria listed in **Table 1** were eligible for inclusion in the review. As the focus of this SLR was on outcomes captured during Omicron BA.2 and BA.5 predominance periods, only papers reporting these subvariants are included here.

**Table 1.**
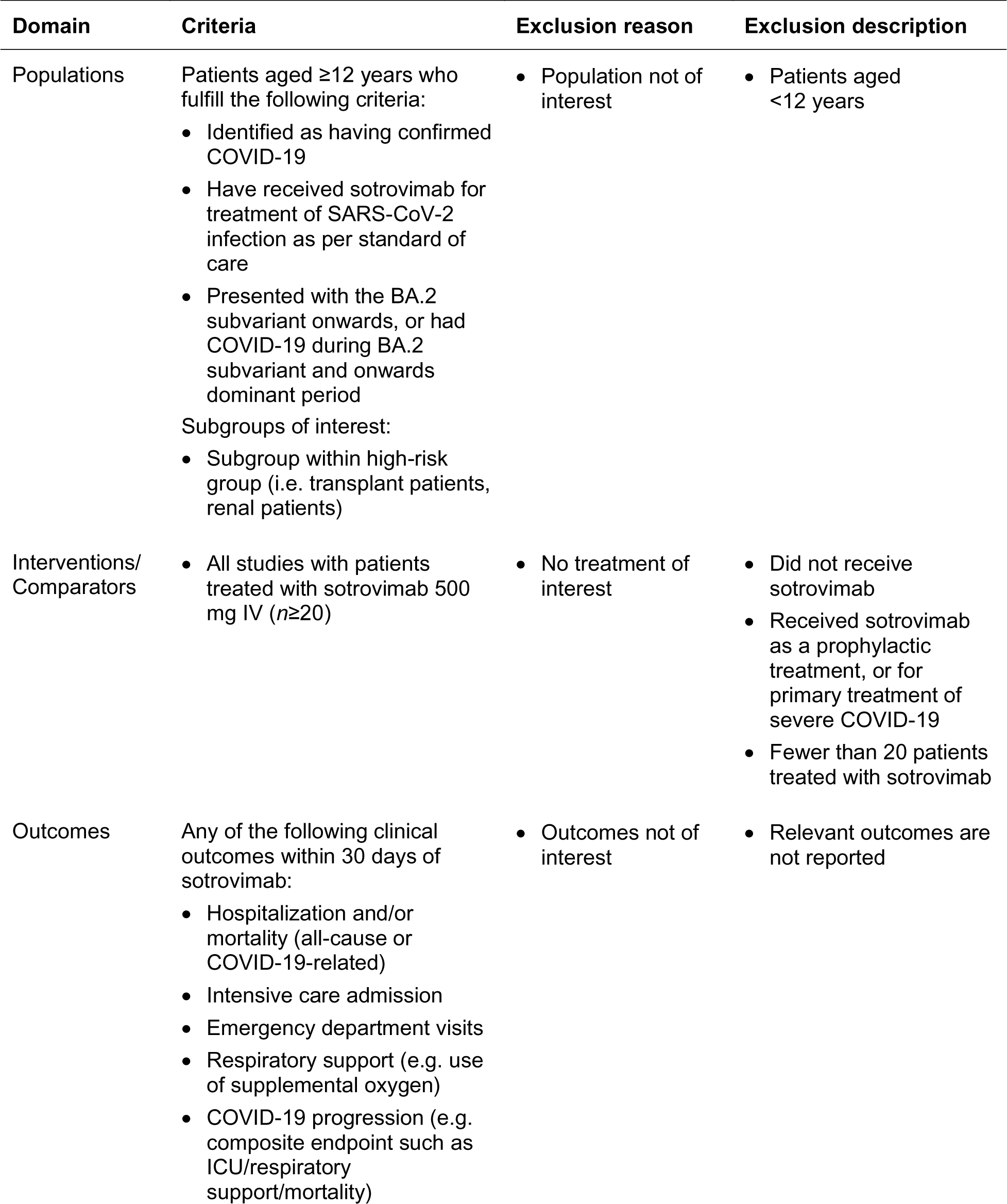
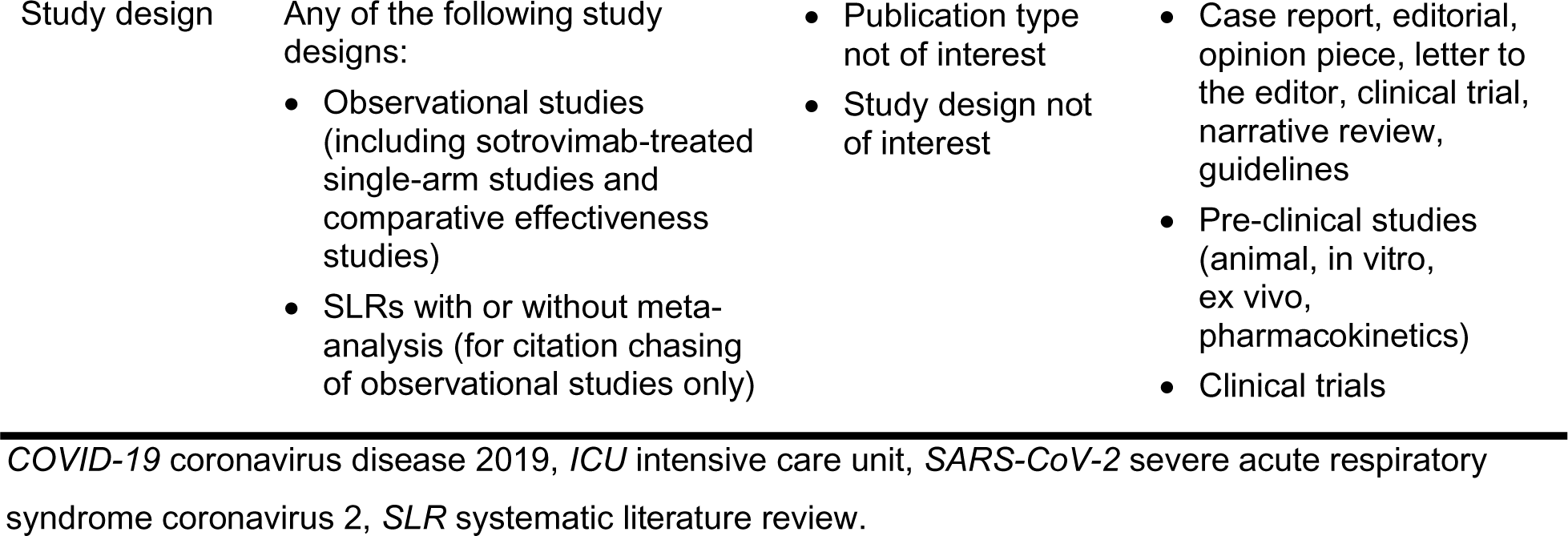
Inclusion and exclusion criteria.

Two independent reviewers evaluated each title and abstract against the defined selection criteria to determine suitability for the SLR, with disagreements resolved by a third reviewer. The same process was applied for the review of the full-text articles.

### Data extraction and quality assessment

Extraction of data from the included studies was performed by a single extractor using a data extraction file designed in Microsoft Excel. An independent researcher reviewed all extracted fields, with discrepancies resolved by a third reviewer.

Extracted information included the study title and reference, study details and design, country(ies), data source, study population, number of patients, data collection period and associated circulating SARS-CoV-2 variants, follow-up duration, sponsor, key baseline characteristics, and clinical outcomes. Clinical outcomes included hospitalization and/or mortality, intensive care admission, emergency department visits, respiratory support (e.g. use of supplemental oxygen), and COVID-19 progression (e.g. composite endpoint such as intensive care unit [ICU]/respiratory support/mortality).

The 8-item Newcastle Ottawa Scale (NOS) was used to assess the quality of each study by considering characteristics that could introduce bias.^24,25^ Studies were assessed based on three broad domains of their design: (1) selection of study groups, (2) comparability of the participants in each group, and (3) ascertainment of either the exposure or outcome of interest for case-control or cohort studies, respectively.^24^ For each study, the maximum attainable score in a NOS quality assessment is 9 (accumulated across all domains), with greater scores representing a lower risk of bias.

## Results

### Study selection

Searches from electronic database and conference abstracts, preprints and citation chasing from relevant SLRs yielded a total of 767 papers (**Figure 1**). After removal of duplicates, 584 unique titles and abstracts were screened, of which 140 were considered admissible for full-text review. Of these, 14 contained clinical outcome data for sotrovimab from the BA.2 and BA.5 periods onwards and were determined eligible for inclusion in the SLR. Reasons for exclusion during the full-text review are detailed in **Figure 1**.

**Figure 1.**
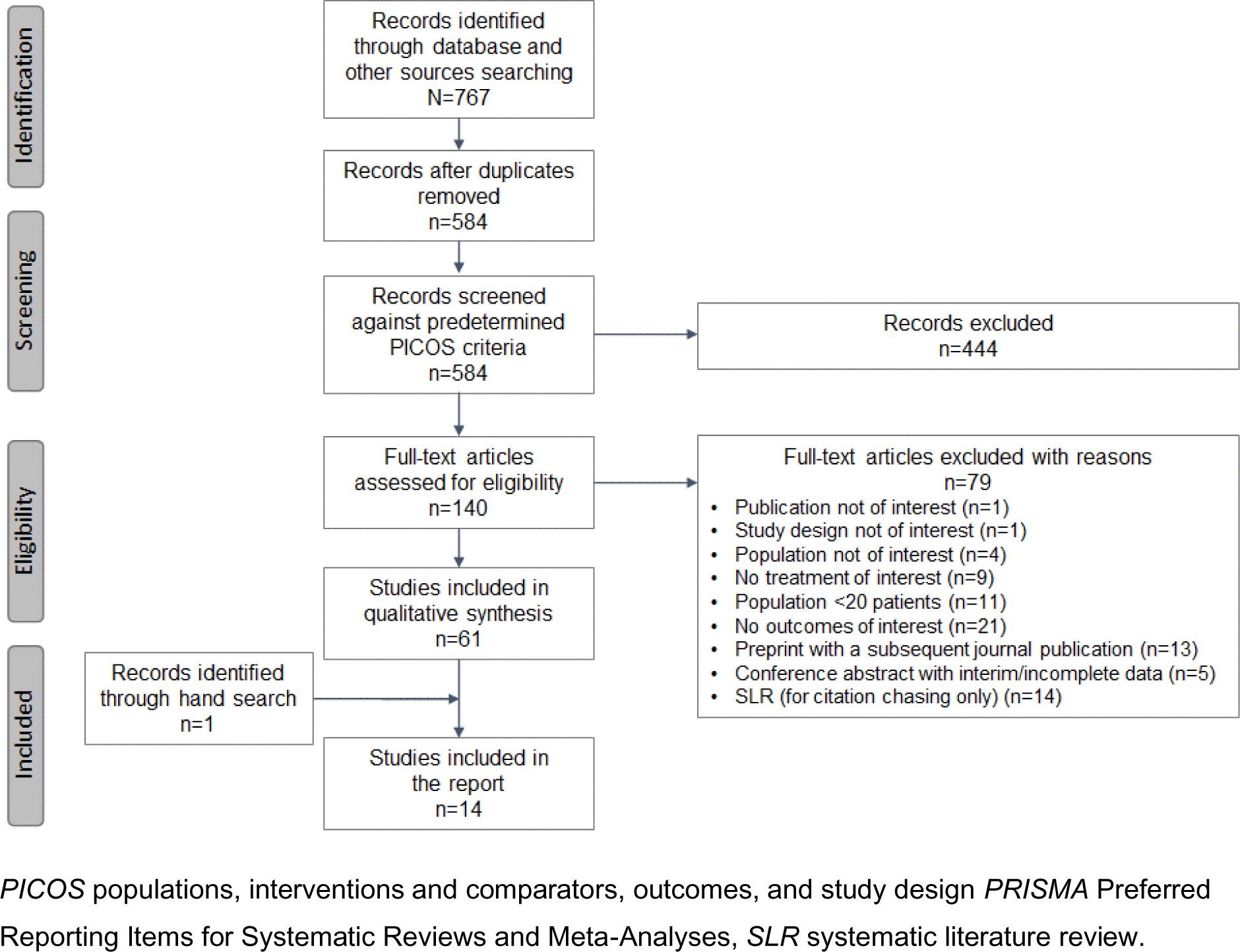
PRISMA flow diagram of studies included in the SLR.

### Study characteristics

An overview of the key characteristics of the 14 observational studies included in the SLR is provided in **Table 2**.

**Table 2.**
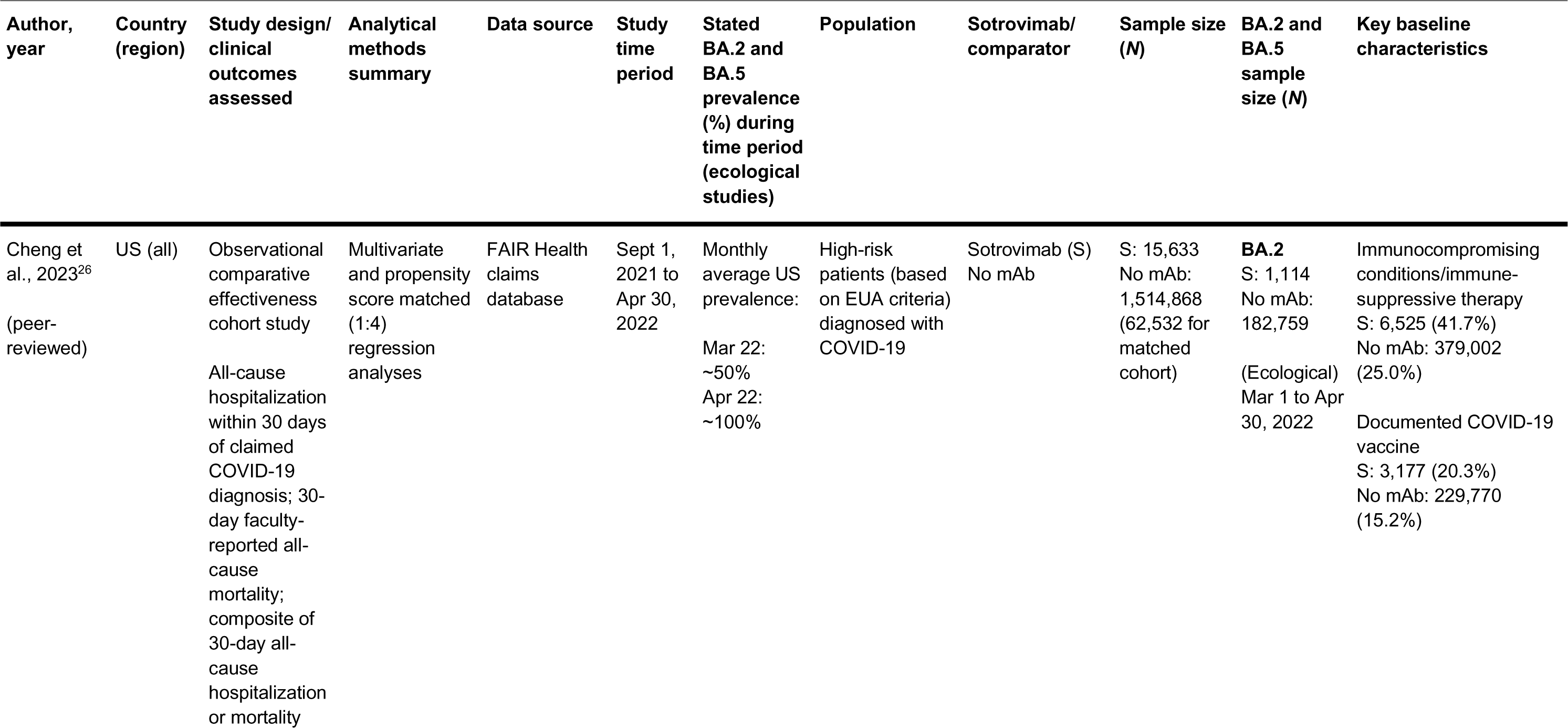
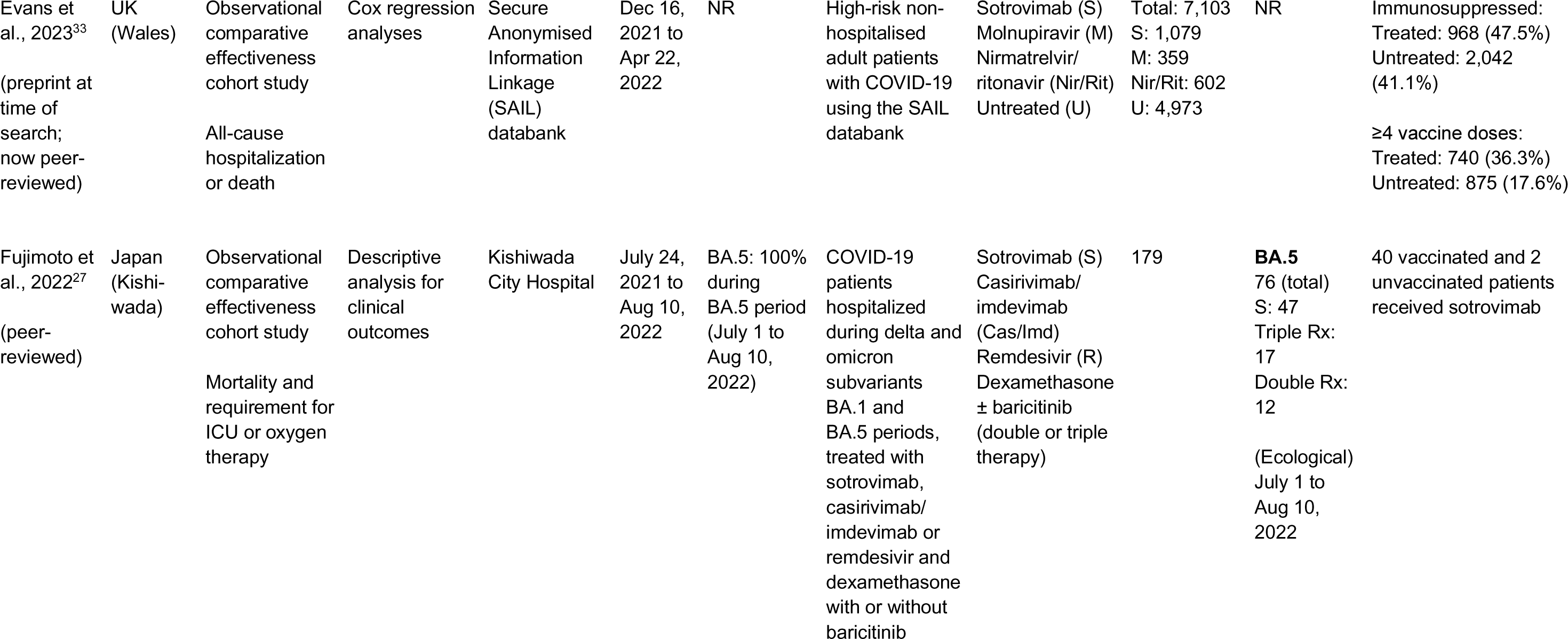
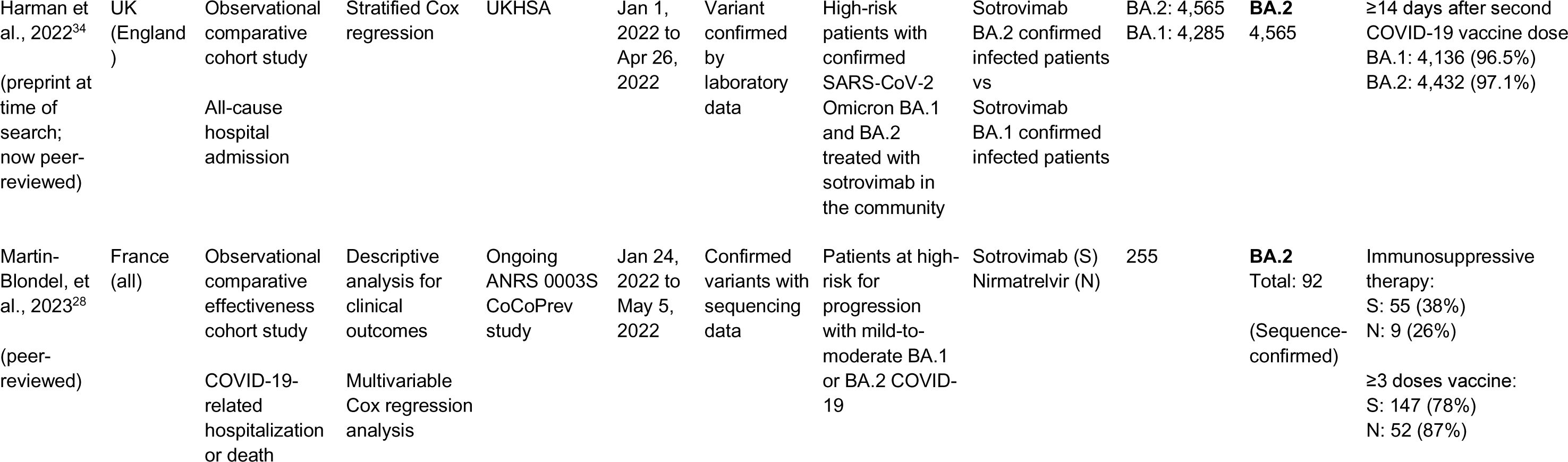
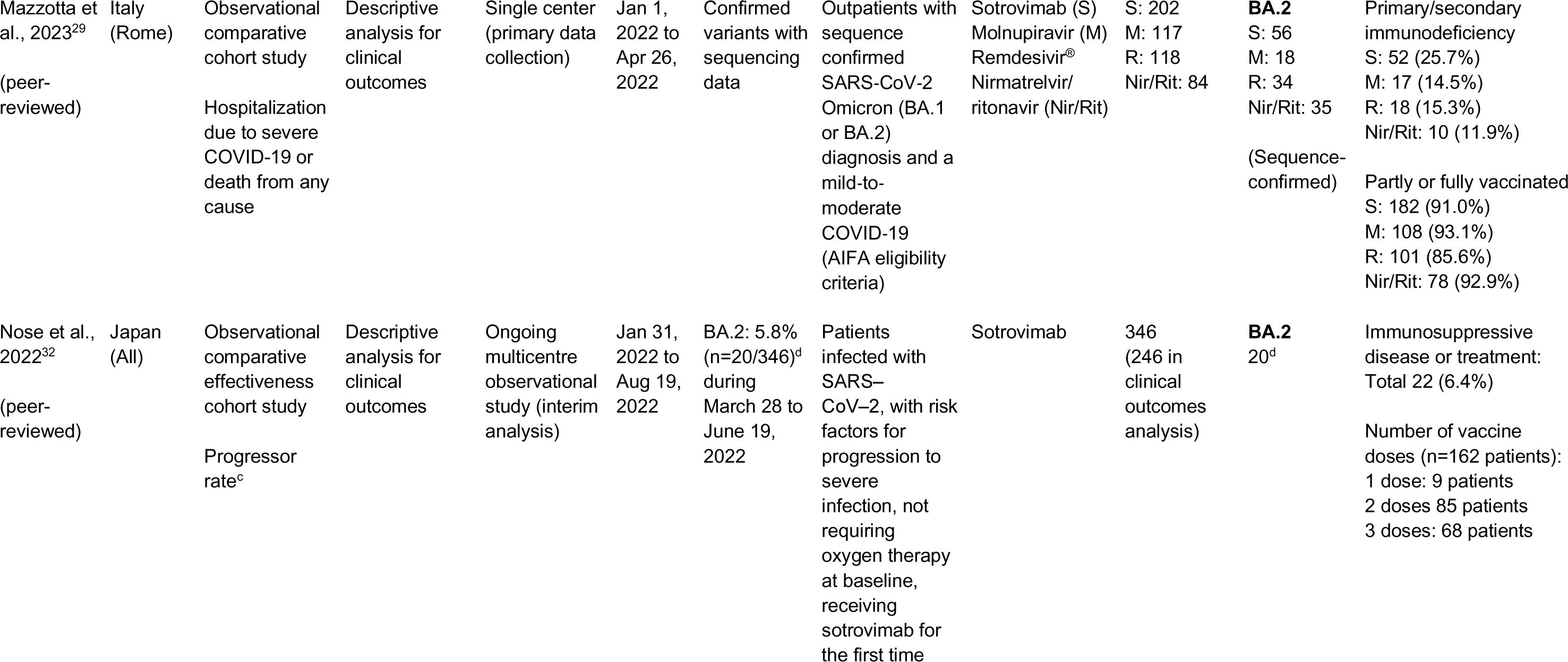
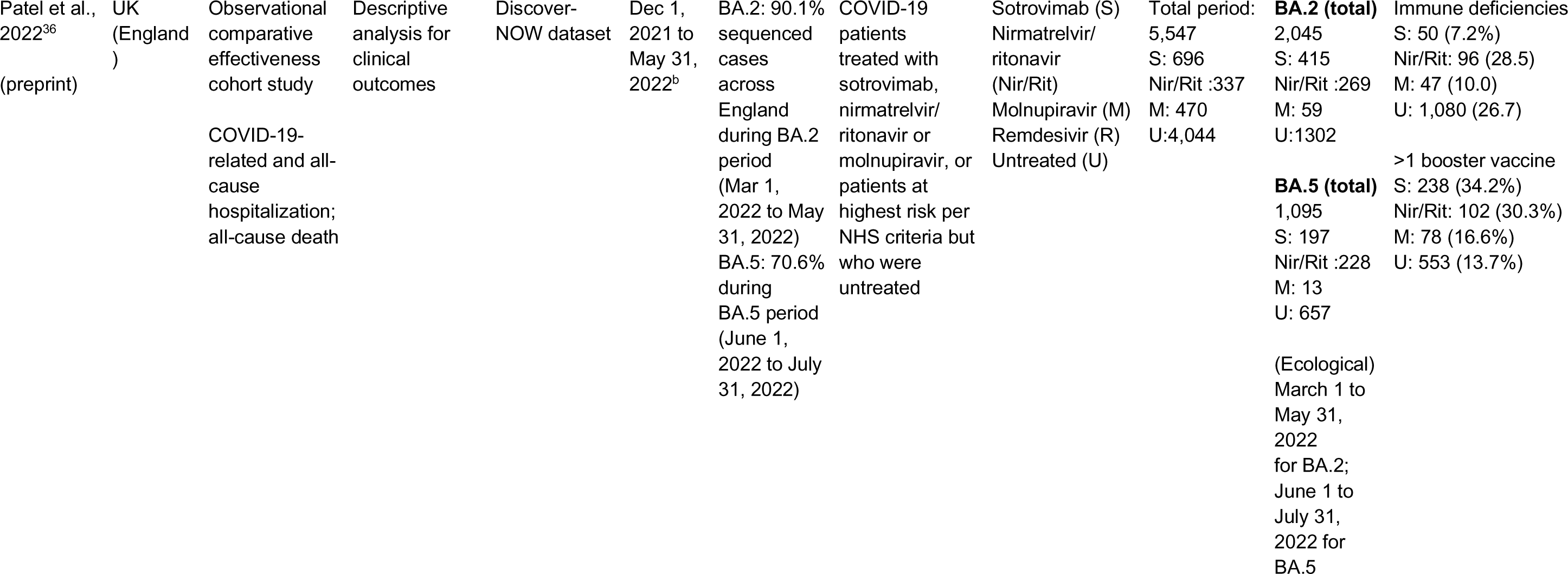
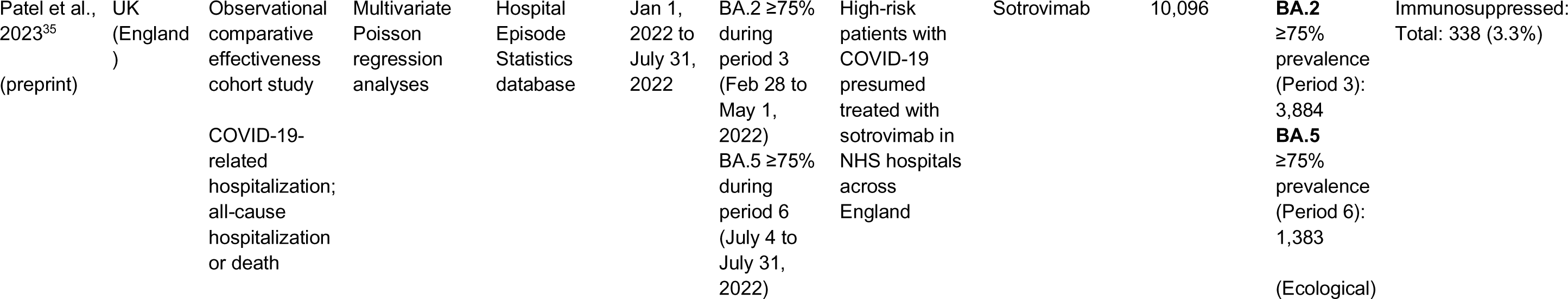
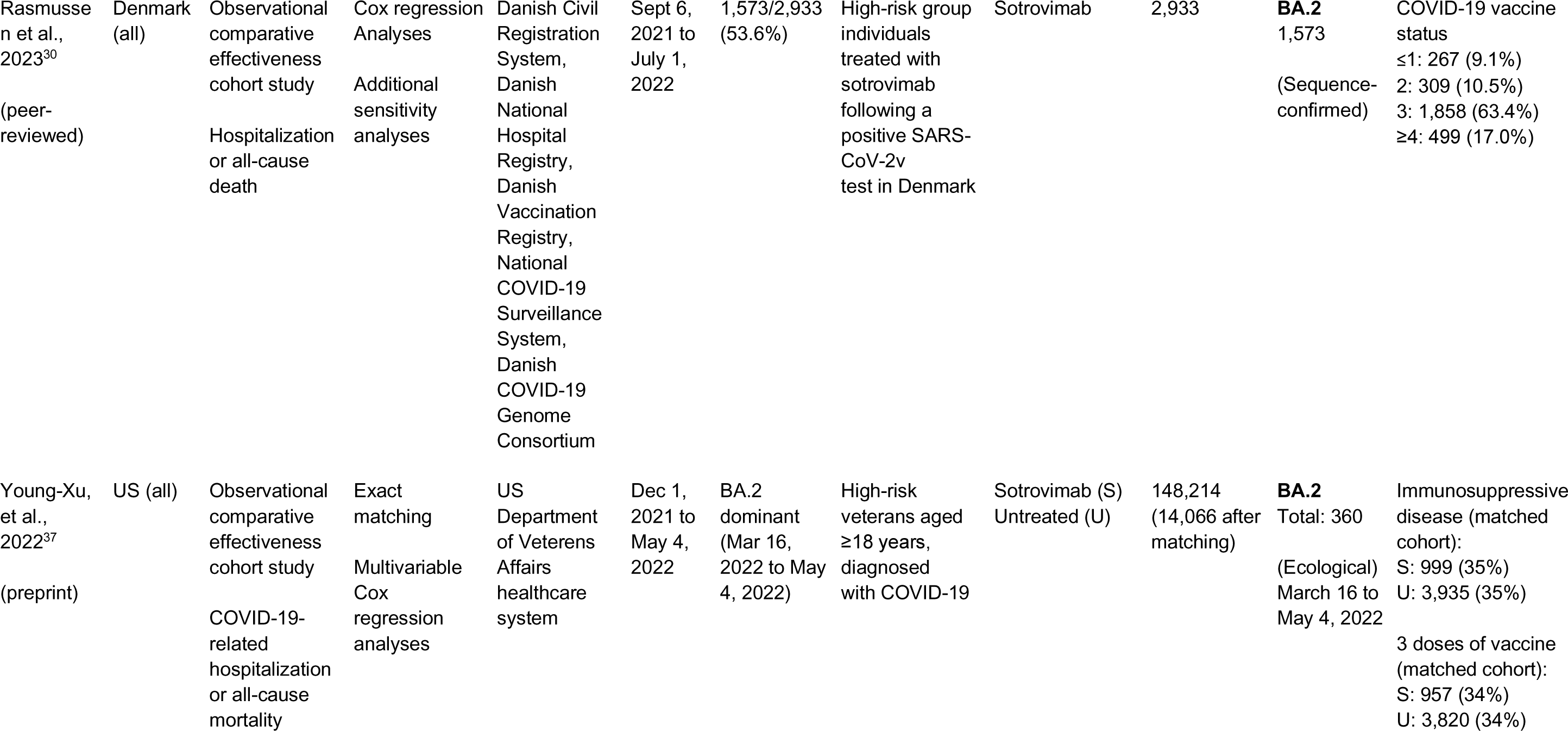
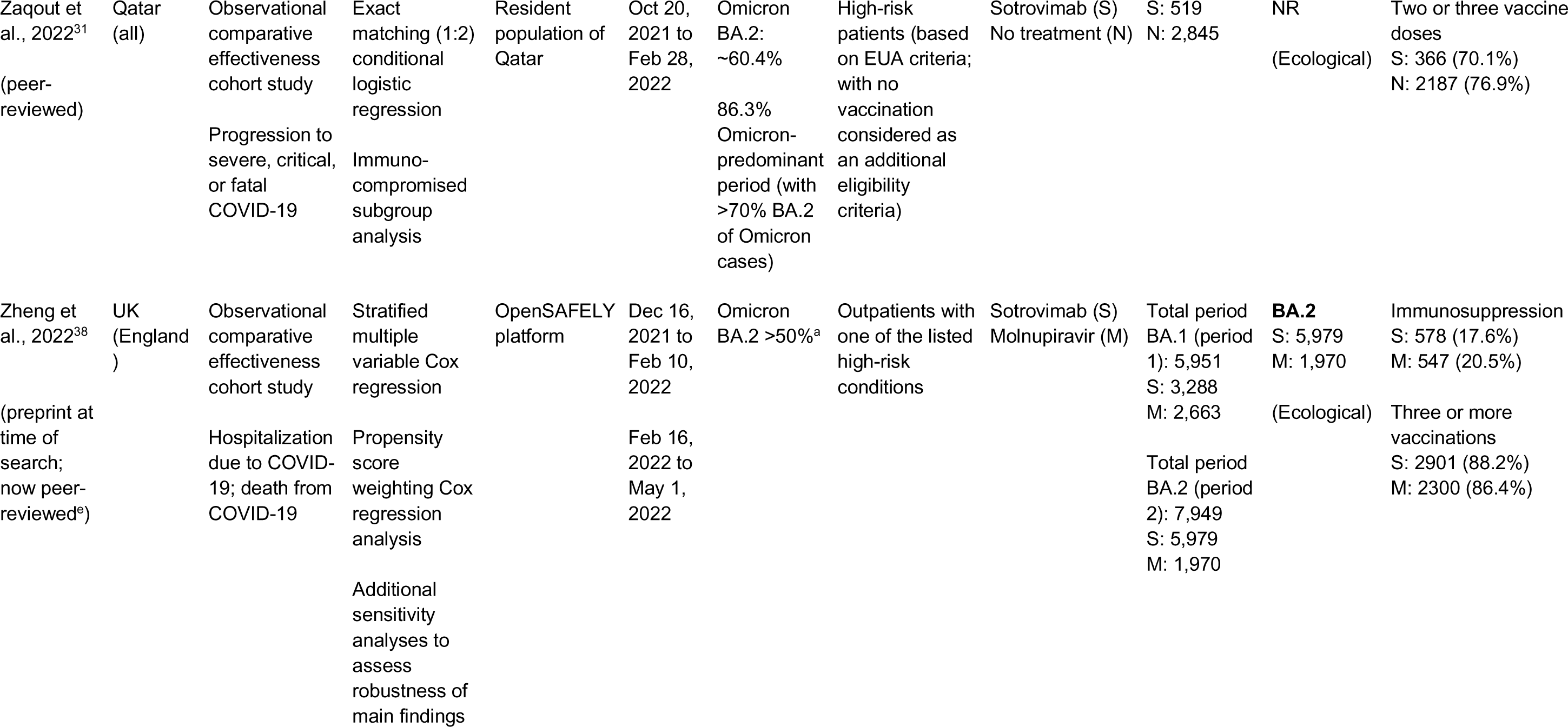
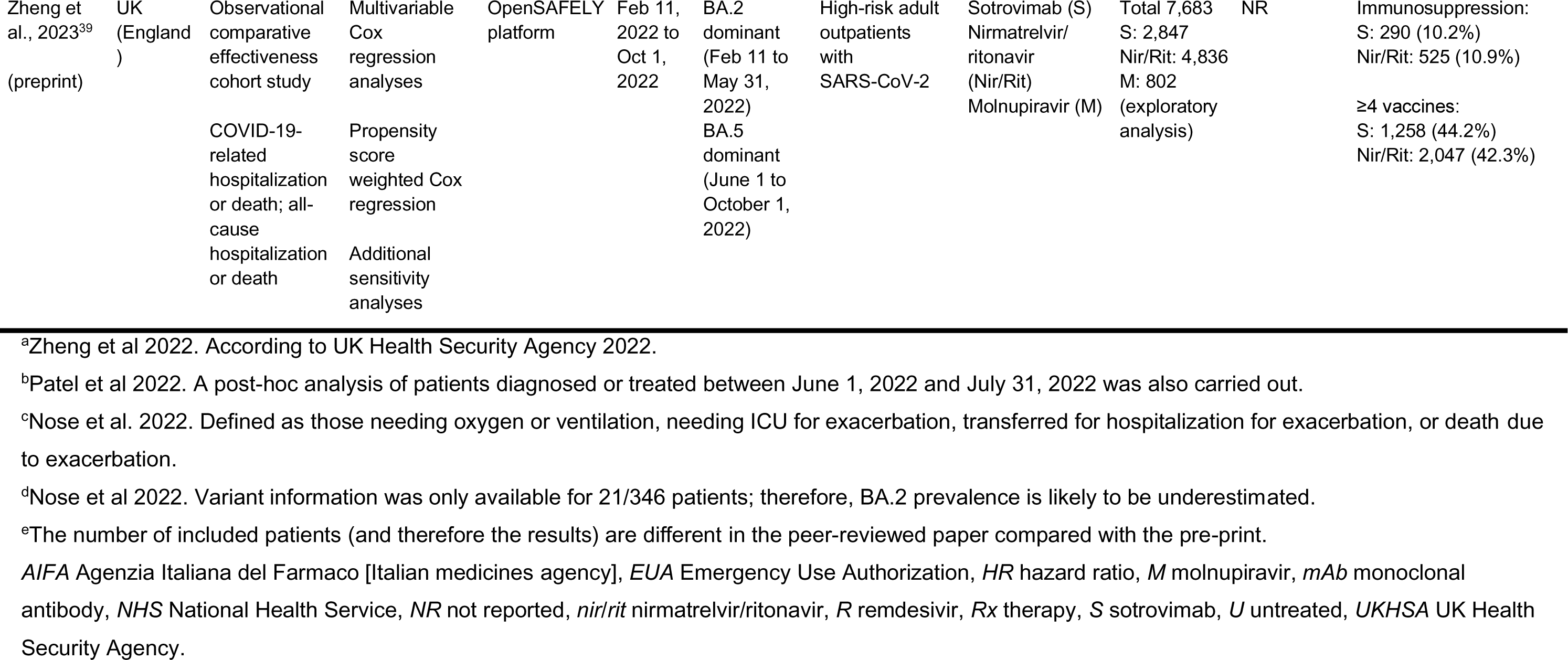
Overview of studies included in the SLR.

Up to February 27, 2023, seven of the 14 studies were published in an international peer-reviewed journal,^26–32^ and seven were published as pre-prints.^33–39^ Three of the preprints have since been published in a peer-reviewed journal.^40–42^ Studies reported on populations from the US (n=2), UK (n=6), Italy (n=1), Denmark (n=1), France (n=1), Japan (n=2), and Qatar (n=1).

Seven studies were conducted via secondary analyses of healthcare data, with sources including OpenSAFELY,^38,39^ Discover-NOW dataset,^36^ SAIL Databank,^33^ and the Hospital Episode Statistics database.^35^ Other data sources included patient electronic medical records or charts,^27,28,32,37^ insurance claims,^26^ and laboratory data.^29^

All studies evaluated clinical outcomes associated with sotrovimab use. Four studies compared the effectiveness of sotrovimab relative to untreated control groups or no mAb treatment.^26,31,33, 37^ Two provided comparative effectiveness data for sotrovimab relative to other treatments (e.g., mAbs, antivirals, corticosteroids).^38,39^ Four studies comprised a single-arm treatment design and compared clinical outcomes of sotrovimab-treated patients during BA.2 and/or BA.5 predominant periods versus the BA.1 period.^30,32,34,35^ Descriptive reporting rates of clinical outcomes (e.g. hospitalization) in sotrovimab-treated patients were used in five studies.^27–29,32,36^

As all studies were observational, sotrovimab was utilized as standard of care in accordance with local guidelines. For the studies in the US, UK, Italy, France, Japan and Qatar, sotrovimab 500 mg was the label recommended dose at the time of the study period. We cannot exclude that another dosage was used for the study in Denmark.

Nine studies reported outcomes for sotrovimab during both Omicron BA.1 and BA.2 predominance.^26,29,30,32–34,36–38^ One study reported outcomes during periods of Omicron BA.1, BA.2 and BA.5 predominance,^36^ two studies during periods of Omicron BA.2 and BA.5 predominance,^35,39^ and one Japanese study during periods of Omicron BA.1 and BA.5 predominance.^27^ Of note, Cheng et al also reported clinical outcomes for March and April 2022 when Omicron BA.2 was becoming predominant in the United States, with estimated prevalence of 50% and 100%, respectively.^26^ Zaqout et al only reported outcomes during a period when both Omicron BA.1 and BA.2 were circulating, without differentiating outcomes by subvariant, but during which >70% of incidence cases were estimated to be BA.2 infections.^31^

Eleven of the 14 studies employed an ecological design, with the date or month of COVID-19 diagnosis used as a proxy for the likelihood of an infection being attributable to the prevalent Omicron subvariant circulating in the country/region at the time.^26,27,30–33,35–39^ The other three studies used sequencing data to ascertain the SARS-CoV-2 subvariant of infection.^28,29,34^

Collectively, the 14 studies included over 1.7 million high-risk patients with COVID-19, defined as those with pre-specified comorbid conditions and/or characteristics leading to progression to severe COVID-19 (note that there is a risk of partial study population overlap between observational studies conducted in the same country). Approximately 41,000 patients received sotrovimab as an early treatment for mild-to-moderate COVID-19. Sample size varied between studies, ranging from 179 patients in a single-center study^27^ to 1,530,501 patients from a nationwide US insurance claims database.^26^ Sample sizes of sotrovimab-treated patients within specific variant predominance periods ranged from n=20–5979 during BA.2 and n=76–1383 during BA.5 predominance. The high-risk populations were heterogeneous, reflecting the differing treatment recommendations in each country at the time of study conduct. As sotrovimab was administered as standard of clinical care, the eligibility criteria for being enrolled in a study reflected the guideline recommendations for sotrovimab as an early COVID-19 treatment in individual countries.

Five studies were conducted in adults aged ≥18 years,^28,33,37–39^ eight studies included patients aged ≥12 years,^26,31,35,36,29,30,32,34^ and one study did not report the age of patients.^27^ The reported mean age of sotrovimab-treated patients in the selected studies ranged from 40^31^ to 79^27^ years.

Of the 14 included studies, seven reported on the composite measure of hospitalization or mortality during Omicron BA.2 and BA.5 predominance, either related to COVID-19^29,34,38,39^ and/or all-cause ^26,33,34,39^ (**Table 3**). Three studies reported estimates for mortality alone^27,30,38^ and four studies reported on hospitalization alone.^30,31,35,36^ One study reported on hospitalization or emergency department or urgent care visits,^37^ and one study briefly reported on the need for intensive care during COVID-19 infection.^27^ The Japanese study by Nose et al included a clinical endpoint of proportion of progressors, defined as patients who required oxygen, non-invasive or invasive ventilation, extracorporeal membrane oxygenation, admission to high care unit or ICU, transfer to another hospital, or died from exacerbation of SARS-CoV-2 infection.^32^ In Japan, patients with COVID-19 were routinely hospitalized at the beginning of treatment. This may explain why the studies by Fujimoto et al^27^ and Nose et al^32^ did not report hospitalization rates.

**Table 3.**
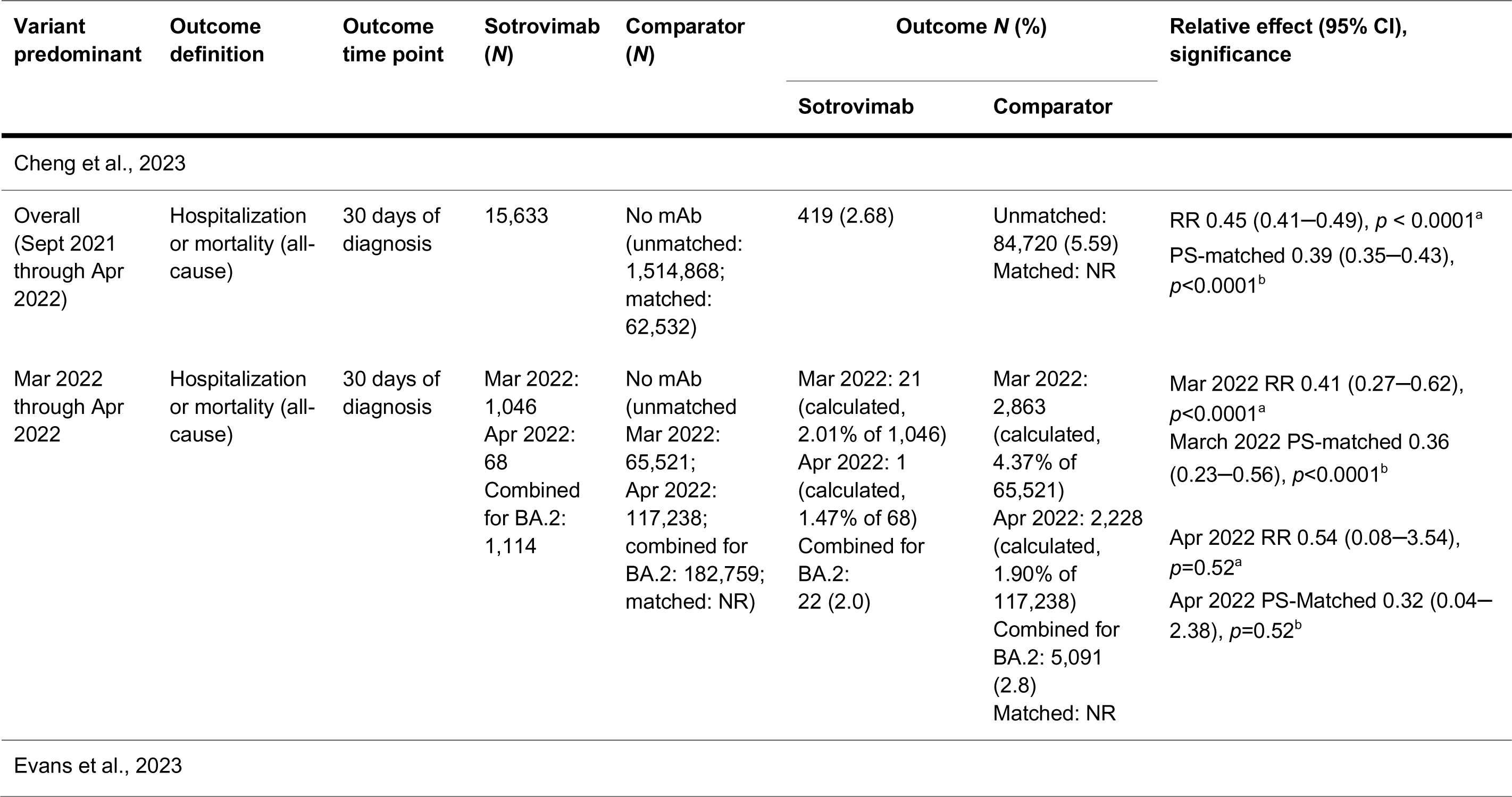
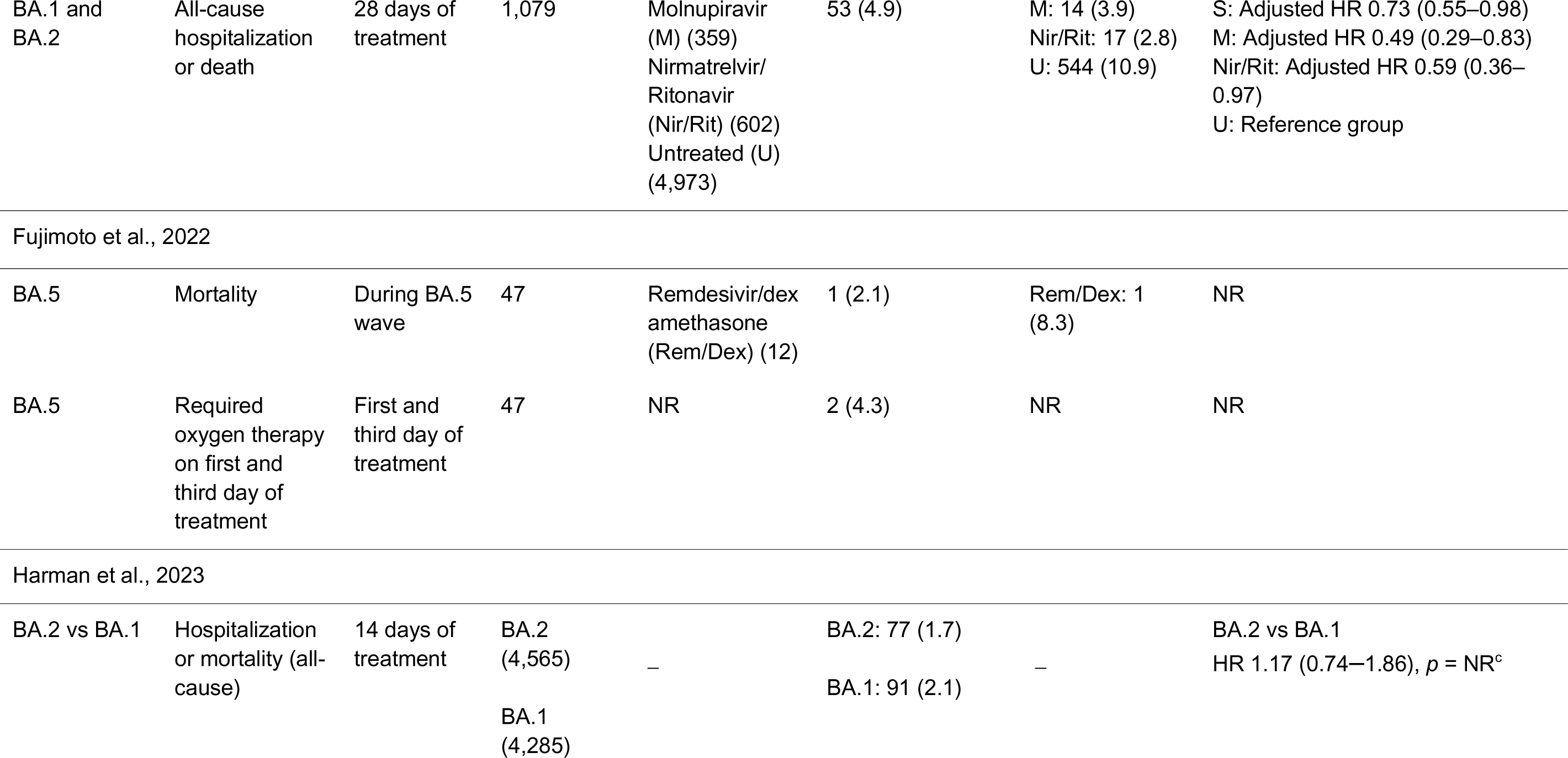
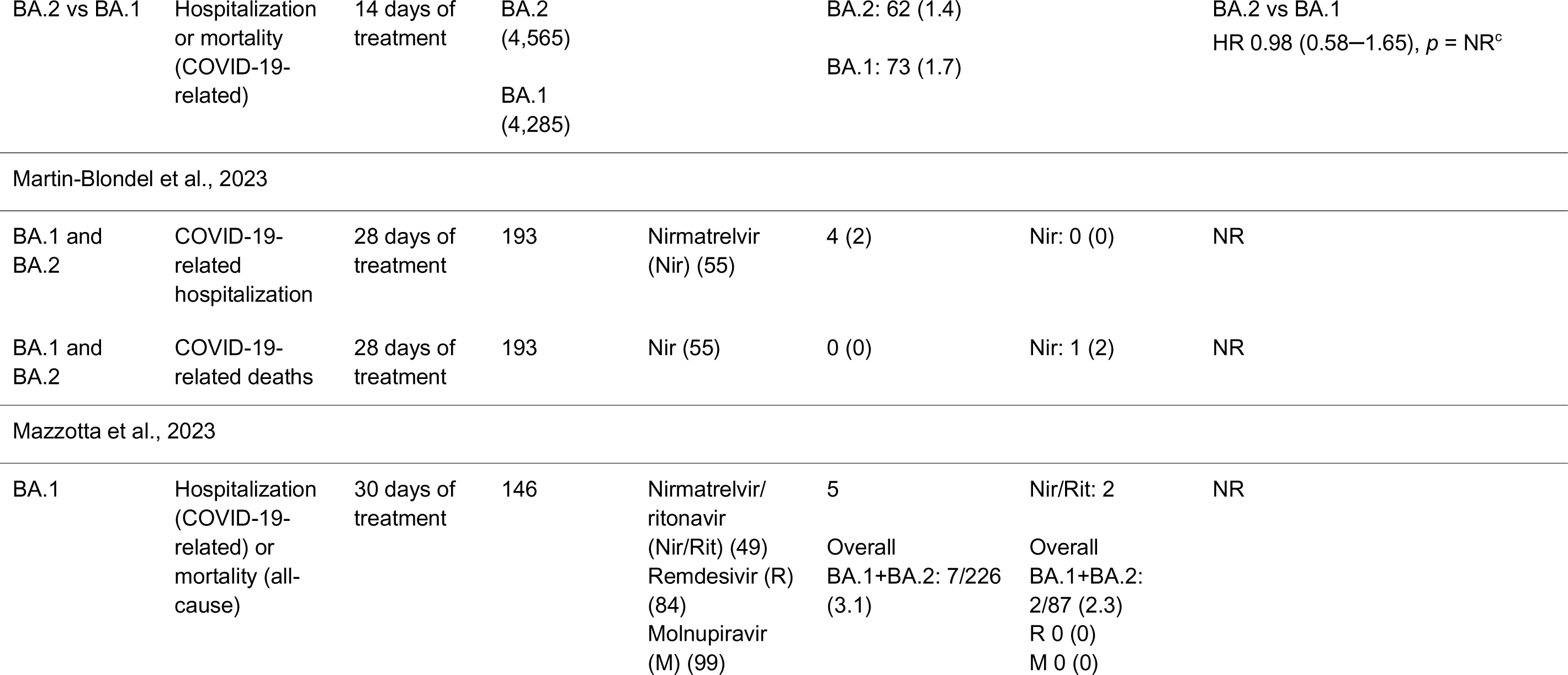
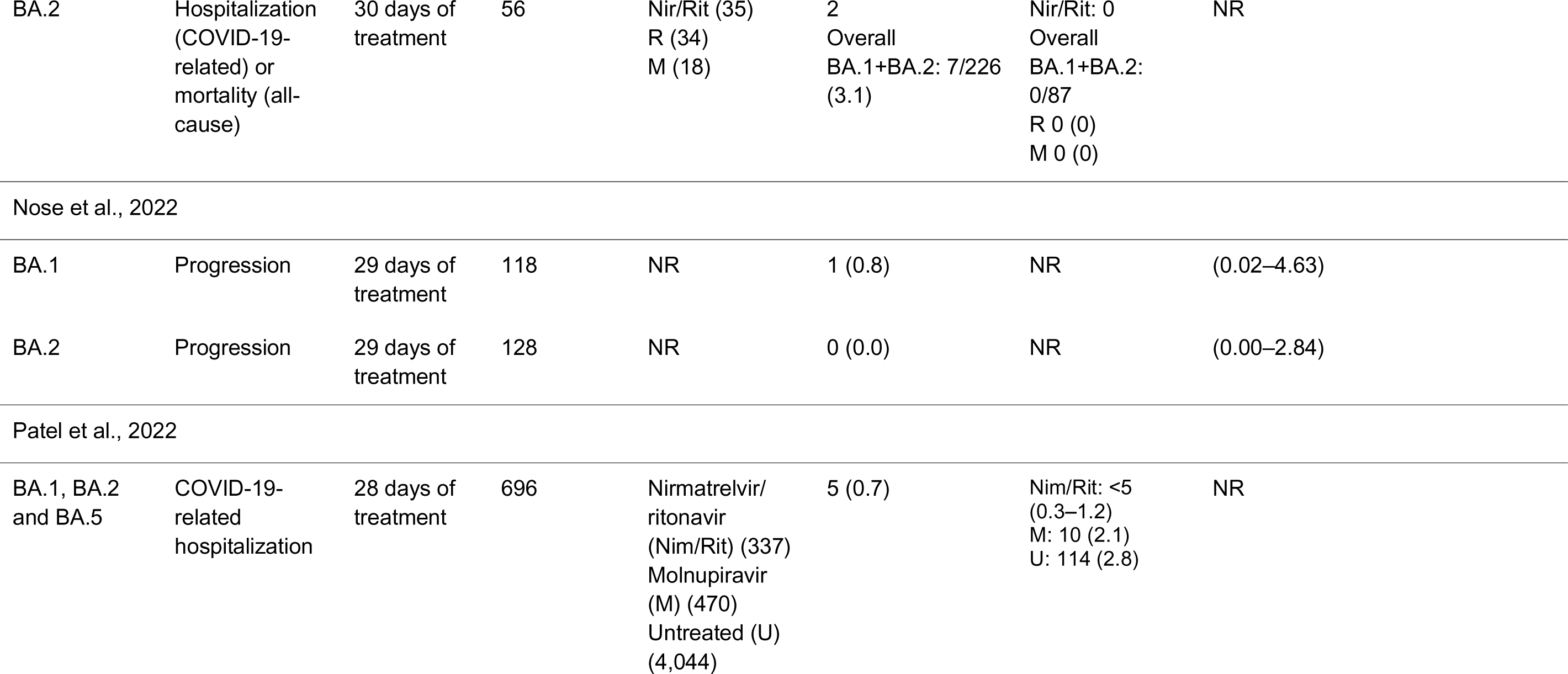
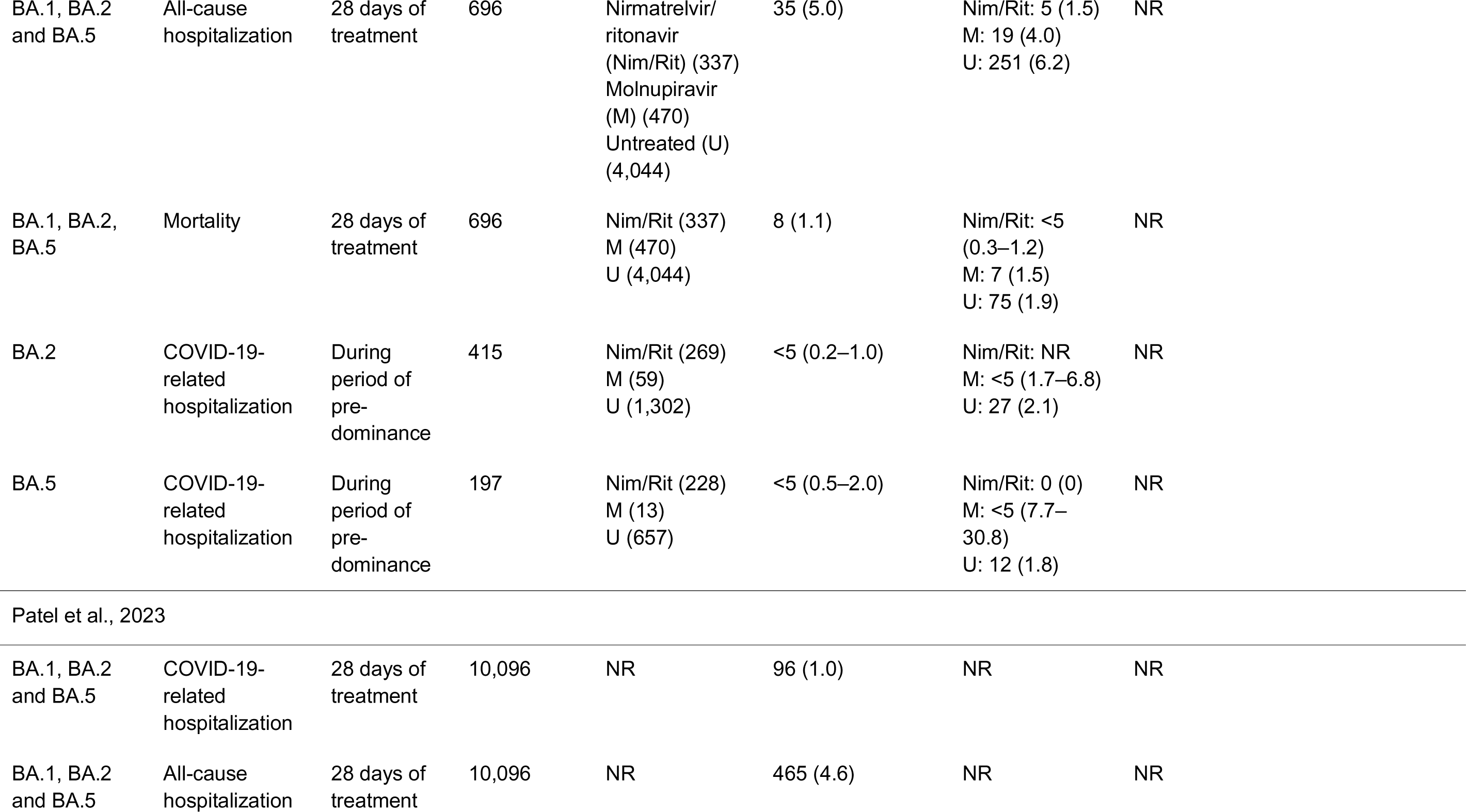
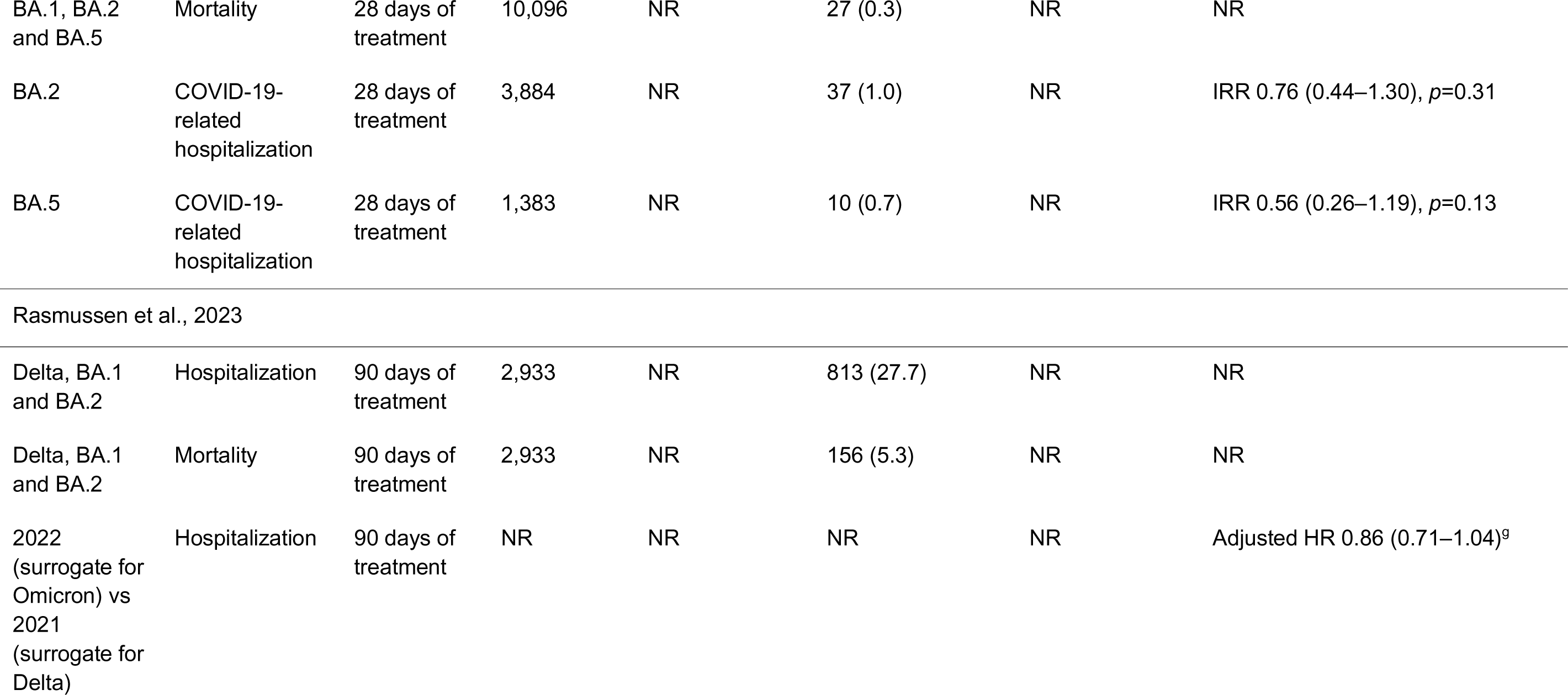
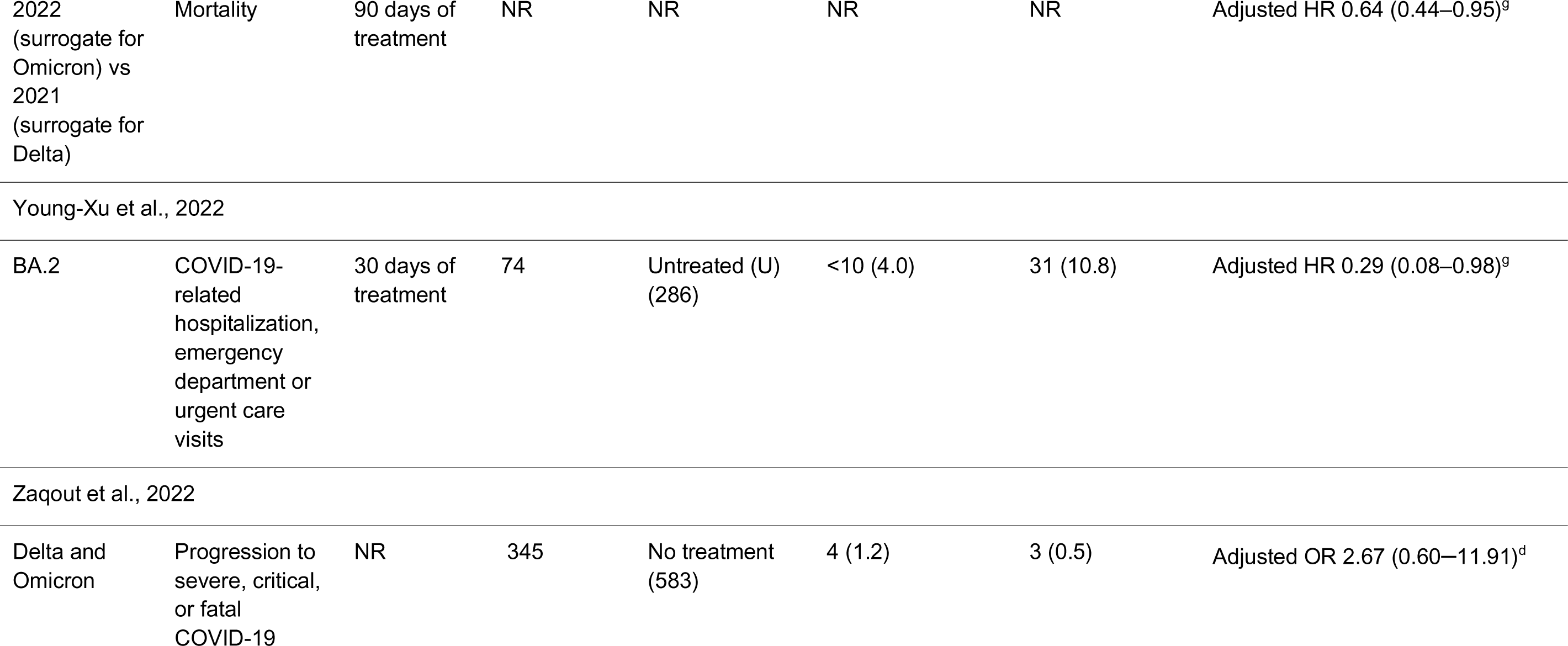
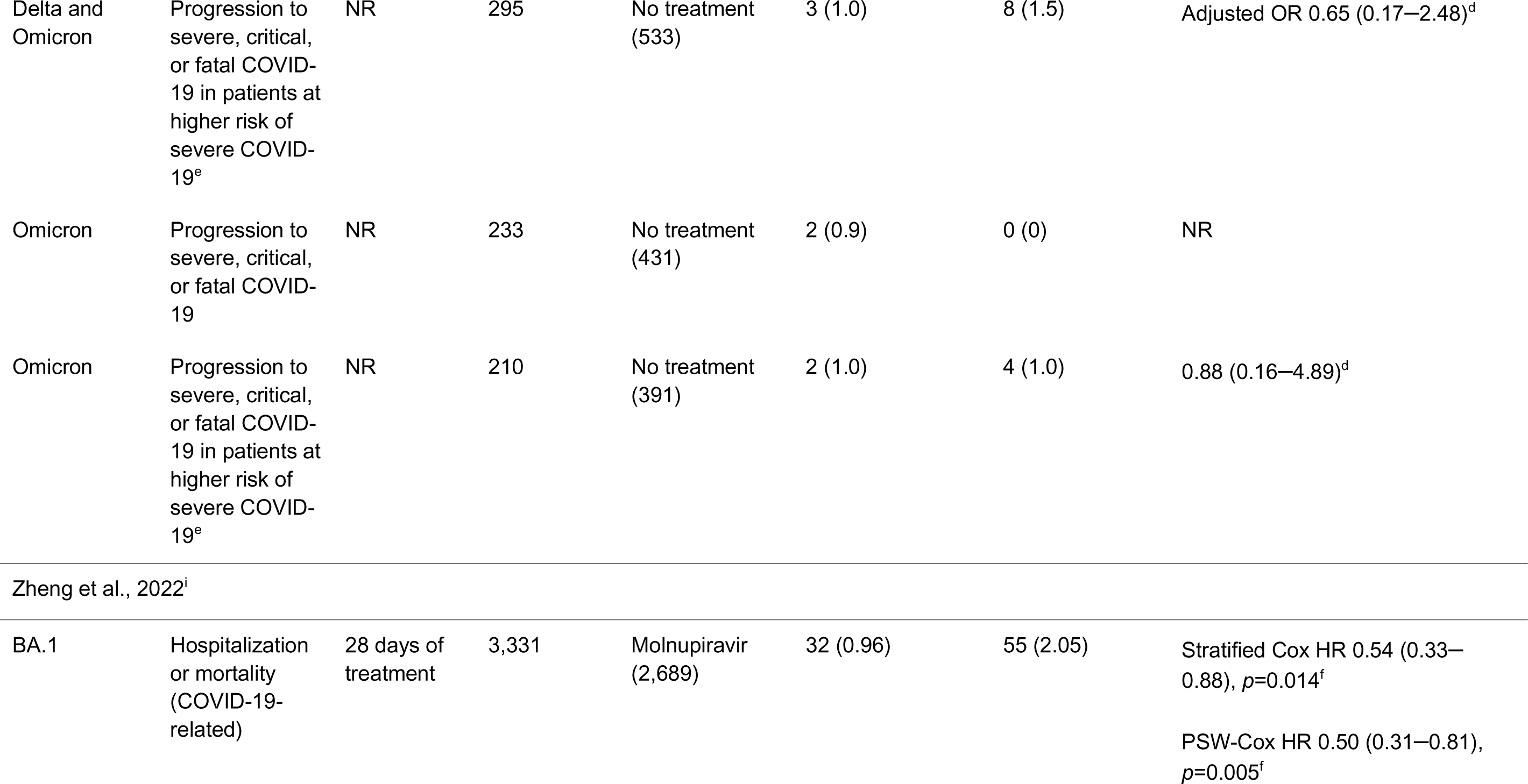
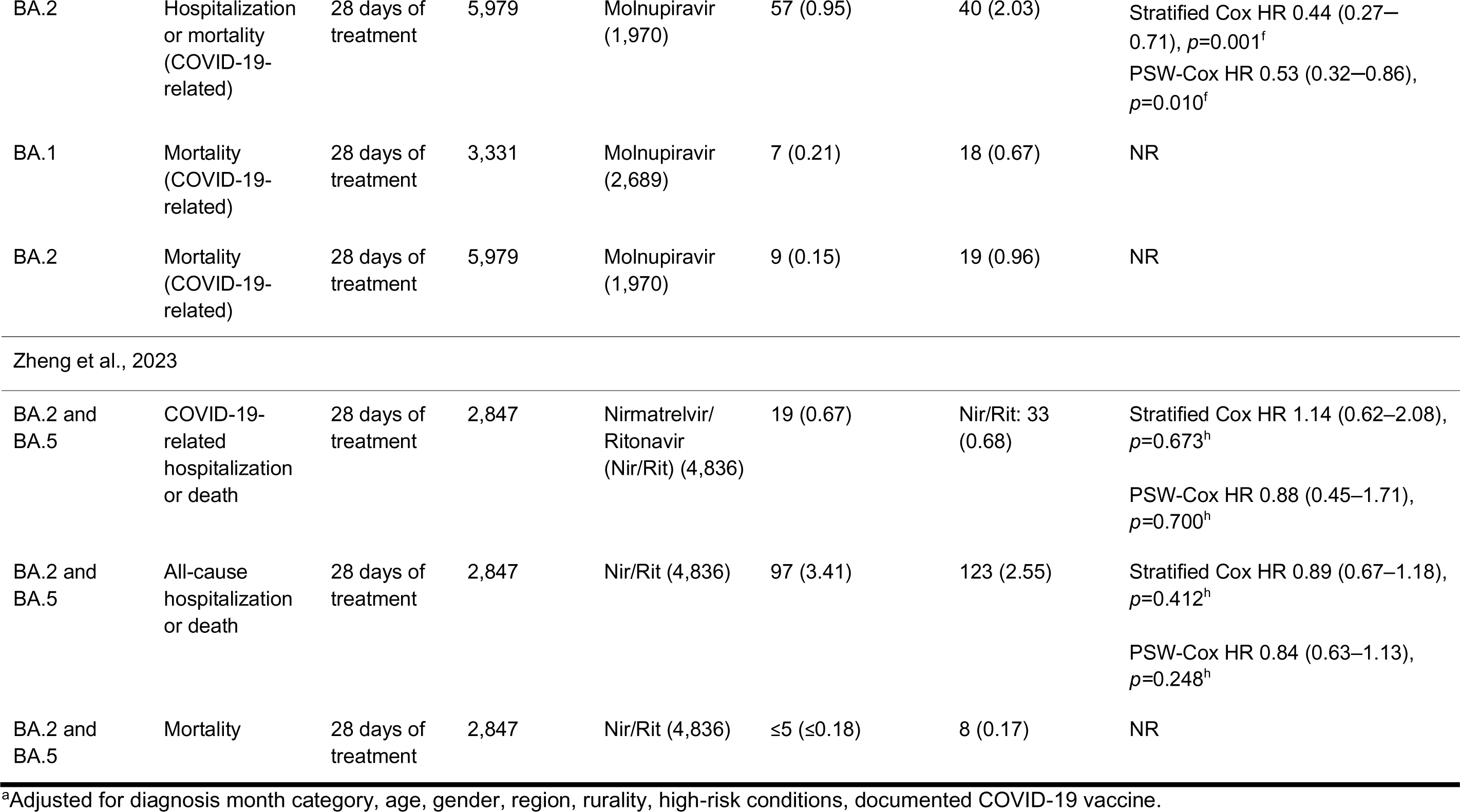
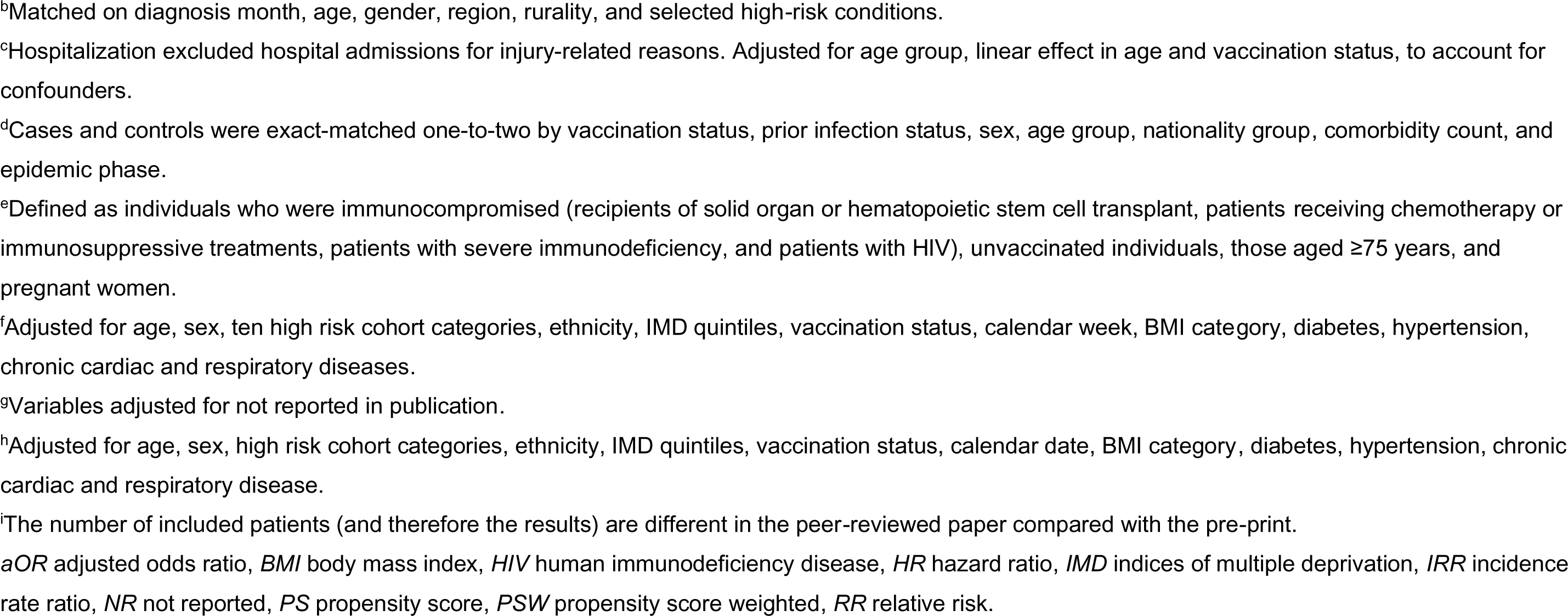
Clinical effectiveness of sotrovimab during Omicron BA.2 and BA.5 predominance.

| Variant predominant | Outcome definition | Outcome time point | Sotrovimab (N) | Comparator (N) | Outcome N (%) |  | Relative effect (95% CI), significance |
| --- | --- | --- | --- | --- | --- | --- | --- |
|  |  |  |  |  | Sotrovimab | Comparator |  |
| Cheng et al., 2023 |  |  |  |  |  |  |  |
| Overall (Sept 2021 through Apr 2022) | Hospitalization or mortality (all-cause) | 30 days of diagnosis | 15,633 | No mAb (unmatched: 1,514,868; matched: 62,532) | 419 (2.68) | Unmatched: 84,720 (5.59)<br>Matched: NR | RR 0.45 (0.41–0.49), *p* < 0.0001<sup>a</sup><br>PS-matched 0.39 (0.35–0.43), *p*<0.0001<sup>b</sup> |
| Mar 2022 through Apr 2022 | Hospitalization or mortality (all-cause) | 30 days of diagnosis | Mar 2022: 1,046<br>Apr 2022: 68<br>Combined for BA.2: 1,114 | No mAb (unmatched Mar 2022: 65,521; Apr 2022: 117,238; combined for BA.2: 182,759; matched: NR) | Mar 2022: 21 (calculated, 2.01% of 1,046)<br>Apr 2022: 1 (calculated, 1.47% of 68)<br>Combined for BA.2: 22 (2.0) | Mar 2022: 2,863 (calculated, 4.37% of 65,521)<br>Apr 2022: 2,228 (calculated, 1.90% of 117,238)<br>Combined for BA.2: 5,091 (2.8)<br>Matched: NR | Mar 2022 RR 0.41 (0.27–0.62), *p*<0.0001<sup>a</sup><br>March 2022 PS-matched 0.36 (0.23–0.56), *p*<0.0001<sup>b</sup><br>Apr 2022 RR 0.54 (0.08–3.54), *p*=0.52<sup>a</sup><br>Apr 2022 PS-Matched 0.32 (0.04–2.38), *p*=0.52<sup>b</sup> |
| Evans et al., 2023 |  |  |  |  |  |  |  |
|  |  |  |  |  | Sotrovimab | Comparator |  |
| BA.1 and BA.2 | All-cause hospitalization or death | 28 days of treatment | 1,079 | Molnupiravir (M) (359)<br>Nirmatrelvir/Ritonavir (Nir/Rit) (602)<br>Untreated (U) (4,973) | 53 (4.9) | M: 14 (3.9)<br>Nir/Rit: 17 (2.8)<br>U: 544 (10.9) | S: Adjusted HR 0.73 (0.55–0.98)<br>M: Adjusted HR 0.49 (0.29–0.83)<br>Nir/Rit: Adjusted HR 0.59 (0.36–0.97)<br>U: Reference group |
| Fujimoto et al., 2022 |  |  |  |  |  |  |  |
| BA.5 | Mortality | During BA.5 wave | 47 | Remdesivir/dexamethasone (Rem/Dex) (12) | 1 (2.1) | Rem/Dex: 1 (8.3) | NR |
| BA.5 | Required oxygen therapy on first and third day of treatment | First and third day of treatment | 47 | NR | 2 (4.3) | NR | NR |
| Harman et al., 2023 |  |  |  |  |  |  |  |
| BA.2 vs BA.1 | Hospitalization or mortality (all-cause) | 14 days of treatment | BA.2 (4,565)<br>BA.1 (4,285) | — | BA.2: 77 (1.7)<br>BA.1: 91 (2.1) | — | BA.2 vs BA.1<br>HR 1.17 (0.74–1.86), $p = \text{NR}^c$ |
|  |  |  |  |  | Sotrovimab | Comparator |  |
| BA.2 vs BA.1 | Hospitalization or mortality (COVID-19-related) | 14 days of treatment | BA.2 (4,565)<br>BA.1 (4,285) |  | BA.2: 62 (1.4)<br>BA.1: 73 (1.7) |  | BA.2 vs BA.1<br>HR 0.98 (0.58–1.65), *p* = NR<sup>c</sup> |
| Martin-Blondel et al., 2023 |  |  |  |  |  |  |  |
| BA.1 and BA.2 | COVID-19-related hospitalization | 28 days of treatment | 193 | Nirmatrelvir (Nir) (55) | 4 (2) | Nir: 0 (0) | NR |
| BA.1 and BA.2 | COVID-19-related deaths | 28 days of treatment | 193 | Nir (55) | 0 (0) | Nir: 1 (2) | NR |
| Mazzotta et al., 2023 |  |  |  |  |  |  |  |
| BA.1 | Hospitalization (COVID-19-related) or mortality (all-cause) | 30 days of treatment | 146 | Nirmatrelvir/ritonavir (Nir/Rit) (49)<br>Remdesivir (R) (84)<br>Molnupiravir (M) (99) | 5<br>Overall<br>BA.1+BA.2: 7/226 (3.1) | Nir/Rit: 2<br>Overall<br>BA.1+BA.2: 2/87 (2.3)<br>R 0 (0)<br>M 0 (0) | NR |
|  |  |  |  |  | Sotrovimab | Comparator |  |
| BA.2 | Hospitalization (COVID-19-related) or mortality (all-cause) | 30 days of treatment | 56 | Nir/Rit (35)<br>R (34)<br>M (18) | 2<br>Overall<br>BA.1+BA.2: 7/226 (3.1) | Nir/Rit: 0<br>Overall<br>BA.1+BA.2: 0/87<br>R 0 (0)<br>M 0 (0) | NR |
| Nose et al., 2022 |  |  |  |  |  |  |  |
| BA.1 | Progression | 29 days of treatment | 118 | NR | 1 (0.8) | NR | (0.02–4.63) |
| BA.2 | Progression | 29 days of treatment | 128 | NR | 0 (0.0) | NR | (0.00–2.84) |
| Patel et al., 2022 |  |  |  |  |  |  |  |
| BA.1, BA.2 and BA.5 | COVID-19-related hospitalization | 28 days of treatment | 696 | Nirmatrelvir/ritonavir (Nim/Rit) (337)<br>Molnupiravir (M) (470)<br>Untreated (U) (4,044) | 5 (0.7) | Nim/Rit: <5 (0.3–1.2)<br>M: 10 (2.1)<br>U: 114 (2.8) | NR |
|  |  |  |  |  | Sotrovimab | Comparator |  |
| BA.1, BA.2 and BA.5 | All-cause hospitalization | 28 days of treatment | 696 | Nirmatrelvir/ritonavir (Nim/Rit) (337)<br>Molnupiravir (M) (470)<br>Untreated (U) (4,044) | 35 (5.0) | Nim/Rit: 5 (1.5)<br>M: 19 (4.0)<br>U: 251 (6.2) | NR |
| BA.1, BA.2, BA.5 | Mortality | 28 days of treatment | 696 | Nim/Rit (337)<br>M (470)<br>U (4,044) | 8 (1.1) | Nim/Rit: <5 (0.3–1.2)<br>M: 7 (1.5)<br>U: 75 (1.9) | NR |
| BA.2 | COVID-19-related hospitalization | During period of pre-dominance | 415 | Nim/Rit (269)<br>M (59)<br>U (1,302) | <5 (0.2–1.0) | Nim/Rit: NR<br>M: <5 (1.7–6.8)<br>U: 27 (2.1) | NR |
| BA.5 | COVID-19-related hospitalization | During period of pre-dominance | 197 | Nim/Rit (228)<br>M (13)<br>U (657) | <5 (0.5–2.0) | Nim/Rit: 0 (0)<br>M: <5 (7.7–30.8)<br>U: 12 (1.8) | NR |
| Patel et al., 2023 |  |  |  |  |  |  |  |
| BA.1, BA.2 and BA.5 | COVID-19-related hospitalization | 28 days of treatment | 10,096 | NR | 96 (1.0) | NR | NR |
| BA.1, BA.2 and BA.5 | All-cause hospitalization | 28 days of treatment | 10,096 | NR | 465 (4.6) | NR | NR |
|  |  |  |  |  | Sotrovimab | Comparator |  |
| BA.1, BA.2 and BA.5 | Mortality | 28 days of treatment | 10,096 | NR | 27 (0.3) | NR | NR |
| BA.2 | COVID-19-related hospitalization | 28 days of treatment | 3,884 | NR | 37 (1.0) | NR | IRR 0.76 (0.44–1.30), $p=0.31$ |
| BA.5 | COVID-19-related hospitalization | 28 days of treatment | 1,383 | NR | 10 (0.7) | NR | IRR 0.56 (0.26–1.19), $p=0.13$ |
| Rasmussen et al., 2023 |  |  |  |  |  |  |  |
| Delta, BA.1 and BA.2 | Hospitalization | 90 days of treatment | 2,933 | NR | 813 (27.7) | NR | NR |
| Delta, BA.1 and BA.2 | Mortality | 90 days of treatment | 2,933 | NR | 156 (5.3) | NR | NR |
| 2022 (surrogate for Omicron) vs 2021 (surrogate for Delta) | Hospitalization | 90 days of treatment | NR | NR | NR | NR | Adjusted HR 0.86 (0.71–1.04)<sup>9</sup> |
|  |  |  |  |  | Sotrovimab | Comparator |  |
| 2022 (surrogate for Omicron) vs 2021 (surrogate for Delta) | Mortality | 90 days of treatment | NR | NR | NR | NR | Adjusted HR 0.64 (0.44–0.95)<sup>g</sup> |
| Young-Xu et al., 2022 |  |  |  |  |  |  |  |
| BA.2 | COVID-19-related hospitalization, emergency department or urgent care visits | 30 days of treatment | 74 | Untreated (U) (286) | <10 (4.0) | 31 (10.8) | Adjusted HR 0.29 (0.08–0.98)<sup>g</sup> |
| Zaqout et al., 2022 |  |  |  |  |  |  |  |
| Delta and Omicron | Progression to severe, critical, or fatal COVID-19 | NR | 345 | No treatment (583) | 4 (1.2) | 3 (0.5) | Adjusted OR 2.67 (0.60–11.91)<sup>d</sup> |
|  |  |  |  |  | Sotrovimab | Comparator |  |
| Delta and Omicron | Progression to severe, critical, or fatal COVID-19 in patients at higher risk of severe COVID-19<sup>e</sup> | NR | 295 | No treatment (533) | 3 (1.0) | 8 (1.5) | Adjusted OR 0.65 (0.17–2.48)<sup>d</sup> |
| Omicron | Progression to severe, critical, or fatal COVID-19 | NR | 233 | No treatment (431) | 2 (0.9) | 0 (0) | NR |
| Omicron | Progression to severe, critical, or fatal COVID-19 in patients at higher risk of severe COVID-19<sup>e</sup> | NR | 210 | No treatment (391) | 2 (1.0) | 4 (1.0) | 0.88 (0.16–4.89)<sup>d</sup> |
| Zheng et al., 2022<sup>i</sup> |  |  |  |  |  |  |  |
| BA.1 | Hospitalization or mortality (COVID-19-related) | 28 days of treatment | 3,331 | Molnupiravir (2,689) | 32 (0.96) | 55 (2.05) | Stratified Cox HR 0.54 (0.33–0.88), $p=0.014^f$<br>PSW-Cox HR 0.50 (0.31–0.81), $p=0.005^f$ |
|  |  |  |  |  | Sotrovimab | Comparator |  |
| BA.2 | Hospitalization or mortality (COVID-19-related) | 28 days of treatment | 5,979 | Molnupiravir (1,970) | 57 (0.95) | 40 (2.03) | Stratified Cox HR 0.44 (0.27–0.71), $p=0.001^f$<br>PSW-Cox HR 0.53 (0.32–0.86), $p=0.010^f$ |
| BA.1 | Mortality (COVID-19-related) | 28 days of treatment | 3,331 | Molnupiravir (2,689) | 7 (0.21) | 18 (0.67) | NR |
| BA.2 | Mortality (COVID-19-related) | 28 days of treatment | 5,979 | Molnupiravir (1,970) | 9 (0.15) | 19 (0.96) | NR |
| Zheng et al., 2023 |  |  |  |  |  |  |  |
| BA.2 and BA.5 | COVID-19-related hospitalization or death | 28 days of treatment | 2,847 | Nirmatrelvir/Ritonavir (Nir/Rit) (4,836) | 19 (0.67) | Nir/Rit: 33 (0.68) | Stratified Cox HR 1.14 (0.62–2.08), $p=0.673^h$<br>PSW-Cox HR 0.88 (0.45–1.71), $p=0.700^h$ |
| BA.2 and BA.5 | All-cause hospitalization or death | 28 days of treatment | 2,847 | Nir/Rit (4,836) | 97 (3.41) | 123 (2.55) | Stratified Cox HR 0.89 (0.67–1.18), $p=0.412^h$<br>PSW-Cox HR 0.84 (0.63–1.13), $p=0.248^h$ |
| BA.2 and BA.5 | Mortality | 28 days of treatment | 2,847 | Nir/Rit (4,836) | $\leq 5$ ($\leq 0.18$) | 8 (0.17) | NR |

<sup>a</sup>Adjusted for diagnosis month category, age, gender, region, rurality, high-risk conditions, documented COVID-19 vaccine.
<sup>b</sup>Matched on diagnosis month, age, gender, region, rurality, and selected high-risk conditions.
<sup>c</sup>Hospitalization excluded hospital admissions for injury-related reasons. Adjusted for age group, linear effect in age and vaccination status, to account for confounders.
<sup>d</sup>Cases and controls were exact-matched one-to-two by vaccination status, prior infection status, sex, age group, nationality group, comorbidity count, and epidemic phase.
<sup>e</sup>Defined as individuals who were immunocompromised (recipients of solid organ or hematopoietic stem cell transplant, patients receiving chemotherapy or immunosuppressive treatments, patients with severe immunodeficiency, and patients with HIV), unvaccinated individuals, those aged $\geq 75$ years, and pregnant women.
<sup>f</sup>Adjusted for age, sex, ten high risk cohort categories, ethnicity, IMD quintiles, vaccination status, calendar week, BMI category, diabetes, hypertension, chronic cardiac and respiratory diseases.
<sup>g</sup>Variables adjusted for not reported in publication.
<sup>h</sup>Adjusted for age, sex, high risk cohort categories, ethnicity, IMD quintiles, vaccination status, calendar date, BMI category, diabetes, hypertension, chronic cardiac and respiratory disease.
<sup>i</sup>The number of included patients (and therefore the results) are different in the peer-reviewed paper compared with the pre-print.
*aOR* adjusted odds ratio, *BMI* body mass index, *HIV* human immunodeficiency disease, *HR* hazard ratio, *IMD* indices of multiple deprivation, *IRR* incidence rate ratio, *NR* not reported, *PS* propensity score, *PSW* propensity score weighted, *RR* relative risk.

Clinical outcomes were generally reported within 28 to 30 days of treatment, with the exception of Harman et al (which reported outcomes within 14 days of treatment^34^) and Rasmussen et al (which reported outcomes within 90 days of COVID-19 diagnosis^30^).

One study (from Qatar) described the results for progression to severe, critical, or fatal COVID-19.^31^ It should be noted that the reasons for COVID-19-related hospital admission in Qatar differed from other included studies. Hospitalization was unrelated to COVID-19 severity and was utilized as a means for dispensing treatment, or as part of a proactive approach to prevent transmission and spread of the disease, as opposed to reducing the risk of further progression.^43^ As such, any comparison of hospitalization rates with the other studies should be considered with caution.

### Quality assessment

Out of the total maximum attainable score of 9 on the NOS, eight studies achieved a score of ≥7, suggesting that they were of comparatively good quality (**Figure 2**).^26,28,30,33,34,37–39^ The remaining studies were awarded a score of 6^29,31,35,36^ or 5.^27,32^ Mazzotta et al was primarily designed to explore changes in SARS-CoV-2 viral load following treatment,^29^ and its score of 6 mainly reflects shortcomings in assessing clinical outcomes rather than overall study quality.

**Figure 2.**
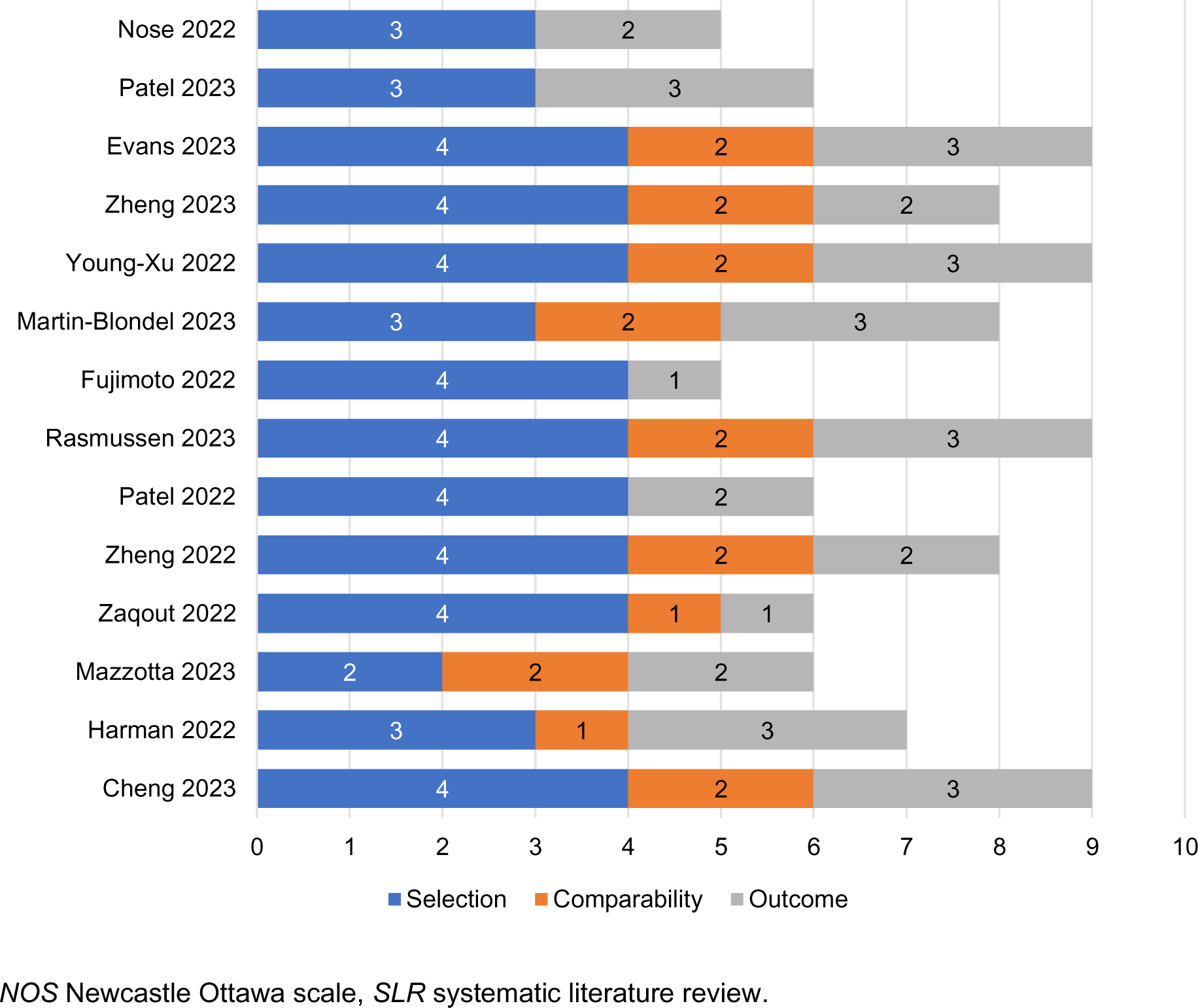
NOS total and bias domain scores across the studies included in the SLR

All studies scored 3 or 4 on the selection bias domain (out of a maximum score of 4), except Mazzotta et al (score of 2), for which the ascertainment of exposure to sotrovimab was not clearly stated.^29^ Most of the studies (n=8/14) scored 2 on the comparability bias domain (out of a maximum score of 2), reporting no major differences in the baseline characteristics of patients or providing adjustment analyses. An exception was Nose et al, which scored zero on this domain due to being a single-arm study.

NOS was not used to assess more specifically the quality of information related to the effectiveness of sotrovimab during Omicron BA.2 or BA.5 predominance. This is of particular relevance to Cheng et al^26^ and Zaqout et al,^31^ which report limited data on Omicron BA.2.

#### Summary of clinical outcomes

The clinical outcomes data extracted from the 14 studies included in this review are provided in **Table 3**.

Rates of COVID-19-related hospitalization or mortality were consistently low across all studies and during periods of Omicron BA.2 and BA.5 predominance (**Table 3**; **Figure 3**). For sotrovimab-treated patients, rates of COVID-19-related hospitalization or death ranged from 0.95%^38^ to 4.0%^37^ during Omicron BA.2 predominance and from 0.5% to 2.0% during BA.5 predominance.^36^

**Figure 3.**
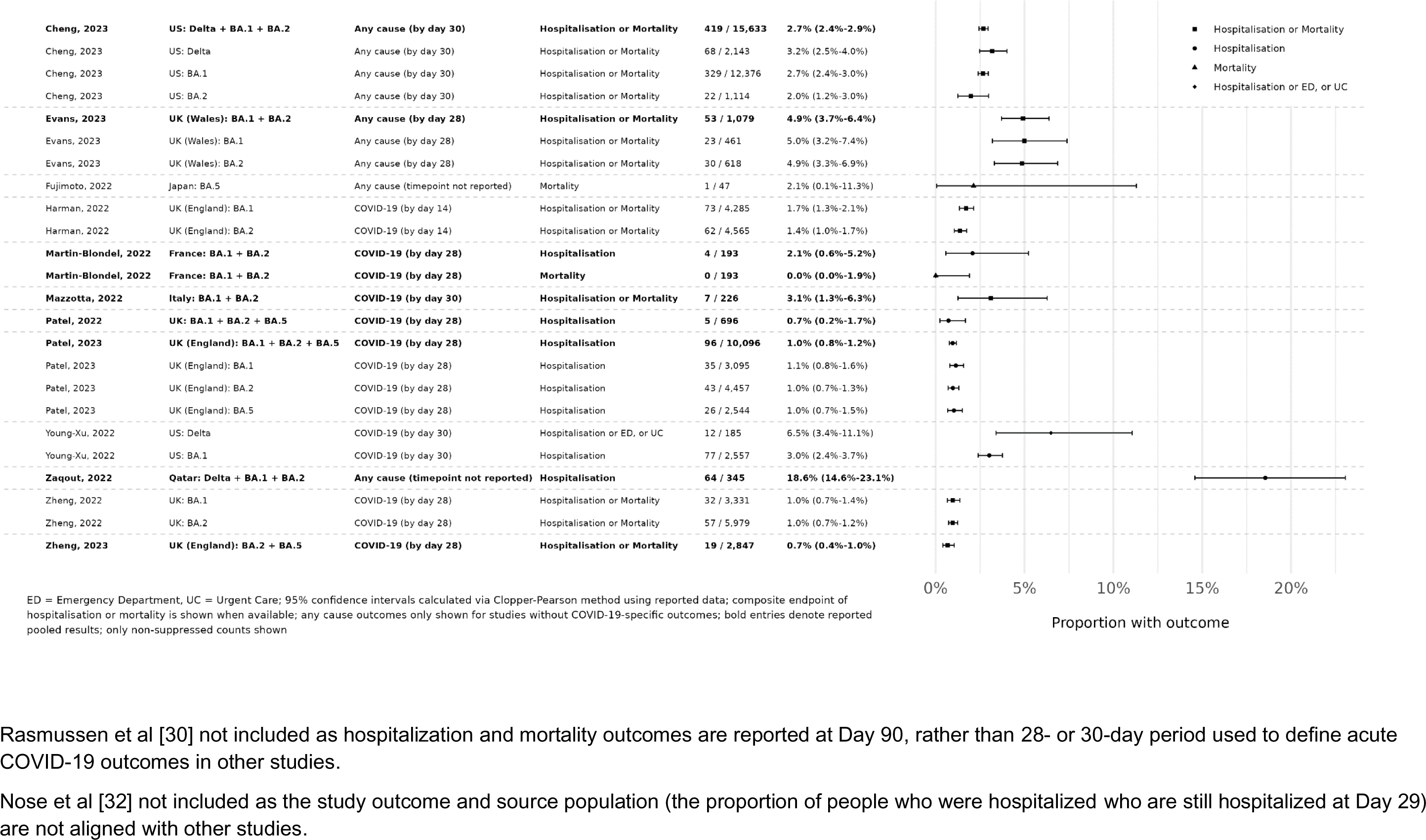
Point estimates for hospitalization or mortality (as a composite endpoint) or clinical progression for sotrovimab-treated patients

The proportions of patients experiencing all-cause hospitalization and/or mortality ranged between 1.7% and 2.0% for the Omicron BA.2 period, as reported by Harman et al (day 14) and Cheng et al (day 30), respectively.^26,33,34^ Only one study (Zheng et al) reported a composite of all-cause hospitalization and/or death in sotrovimab-treated patients during the BA.5 predominance period;^39^ the reported rate (3.4%) was combined with the BA.2 period.^39^

Zheng et al. reported a COVID-19-related mortality rate of 0.15% during Omicron BA.2 predominance for patients treated with sotrovimab (n=9/5979), versus 0.96% for patients treated with the antiviral molnupiravir (n=19/1970).^38^ COVID-19-related mortality during the combined BA.2 and BA.5 predominance periods was estimated at ≤0.18% for the sotrovimab group (n=≤5/2847) vs 0.17% for nirmatrelvir/ritonavir (n=8/4836),^39^ while all-cause mortality during BA.5 predominance was estimated at 2.1% (n=1/47) for the sotrovimab group vs 8.3% (n=1/12) for remdesivir + dexamethasone.^27^

#### Clinical effectiveness of sotrovimab vs control (untreated or no mAb)

Four studies examined the clinical effectiveness of sotrovimab vs a control during Omicron BA.2 predominance.^26,31,33,37^

The US-based study by Cheng et al. reported that sotrovimab was associated with a lower risk of 30-day all-cause hospitalization or mortality compared with no mAb treatment during March and April 2022 (BA.2 period) (**Table 3**).^26^ In March 2022, sotrovimab treatment (n=1046) resulted in a significant reduction in propensity score-matched relative risk (RR) of 64% (adjusted RR 0.36, 95% CI 0.23–0.56; p<0.001) in 30-day all-cause hospitalization or mortality vs patients not treated with a mAb. In April 2022, the propensity score-matched RR reduction was 68% (adjusted RR 0.32, 95% CI 0.04–2.38; p=0.519) compared with patients not treated with a mAb.

The Zaqout et al study in Qatar reported that the overall (periods of Delta and Omicron predominance combined) adjusted odds ratio (OR) of disease progression to severe, critical or fatal COVID-19 for the exact-matched sotrovimab-treated versus untreated control group was 2.67 (95% CI: 0.60–11.91) (**Table 3**).^31^ An adjusted OR of disease progression during the Omicron-dominated time period could not be calculated as none of the 431 untreated patients were observed to have progressed; two of the 233 (0.9%) sotrovimab treated-patients progressed during this phase. In the same study, among patients described as being at higher risk of severe forms of COVID-19 (immunocompromised, unvaccinated individuals, aged ≥75 years, and pregnant women) sotrovimab-treated patients had lower odds of progression compared with untreated patients (adjusted OR 0.65, 95% CI 0.17–2.48). Restricting the analysis to the Omicron-predominant period (December 19, 2021 to February 28, 2022) for the subgroup of higher-risk patients yielded an adjusted OR of 0.88 (95% CI 0.16–4.89) (**Table 3**).

In the US study by Young-Xu et al, treatment with sotrovimab during BA.2 predominance was associated with a reduced risk of COVID-19-related hospitalization, emergency department, or urgent care visits (n=<10/74) within 30 days vs the exact-matched untreated control group (n=31/286; adjusted hazard ratio [HR] 0.29 [95% CI 0.08–0.98]) (**Table 3**).^37^ During the BA.1 period, the adjusted HR of 30-day COVID-19-related hospitalization or all-cause mortality in the sotrovimab group (n=92/2557) vs the group that received no treatment (n=735/10,297) was 0.30 (95% CI 0.23–0.40).

In a UK study by Evans et al, the adjusted HR of all-cause hospitalization or death within 28 days during the study period (BA.1 and BA.2 predominant periods combined) was reported as 0.73 (95% CI 0.55–0.98) for unmatched sotrovimab vs untreated control groups (**Table 3**).^33^

#### Clinical effectiveness of sotrovimab vs active comparators

Compared with molnupiravir, sotrovimab was associated with a lower risk of COVID-19-related hospitalization or death during the BA.2 predominance period in England (February 16 to May 1, 2022), after adjusting for demographics, high-risk cohort categories, vaccination status, calendar time, BMI, and other comorbidities (adjusted HR 0.44, 95% CI 0.27–0.71; p=0.001; propensity score weighted Cox model, adjusted HR 0.53, 95% CI 0.32–0.86, p=0.01).^38^

During the BA.2 (February 11 to May 31, 2022) and BA.5 (June 1 to October 1, 2022) predominance periods in England, treatment with nirmatrelvir/ritonavir was associated with a similar risk of COVID-19-related hospitalization or death to sotrovimab (adjusted HR 1.35, 95% CI 0.54–3.34, and 0.74, 95% CI 0.31–1.78, respectively, using a fully-adjusted stratified Cox model).^39^

#### Comparison of clinical outcomes between periods of different circulating variants

Five studies compared clinical outcomes following sotrovimab treatment during the Omicron BA.1 period and the BA.2 and/or BA.5 predominance periods (**Table 3**).^30,32–35^

In the Harman et al study in England, risk of hospital admission with a length of stay of ≥2 days within 14 days of community treatment with sotrovimab showed no statistically significant difference between BA.1 (2.1%, n=91/4285) and BA.2 (1.7%, n=77/4565) (HR 1.17, 95% CI 0.74–1.86).^34^ Rasmussen et al reported no difference in risk of all-cause mortality and all-cause hospitalization (≥24 hours within 90 days of COVID-19 diagnosis) between Omicron BA.2 (n=1573) and BA.1 (n=381) subvariants in patients in Denmark treated with sotrovimab (adjusted HR 1.04, 95% CI 0.84–1.29 for all-cause hospitalization; adjusted HR 1.04, 95% CI 0.59–1.83 for mortality).^30^ Similarly, in a subanalysis of the study by Evans et al, all-cause hospitalization or death rates among patients in the UK treated with sotrovimab during the BA.1 and BA.2 periods were similar (5.0% vs 4.9%, respectively), with no significant difference between the subvariant time periods (HR 0.76 [95% CI 0.50– 1.18] vs. 0.70 [95% CI 0.48–1.03], respectively).^33^ In another UK study, Patel et al reported no difference in risk of COVID-19-related hospitalization during the Omicron BA.2 (1.0%) and BA.5 (0.7%) predominance periods vs the BA.1 (1.0%) phase among patients treated with sotrovimab [incidence rate ratio (IRR) 0.76, 95% CI 0.44–1.30, p=0.31, and 0.56, 95% CI 0.26–1.19, p=0.13, respectively).^35^ In an interim analysis of a Japanese study, Nose et al reported similar rates of progression for sotrovimab-treated patients infected with Omicron BA.1 (0.8%; n=1/118, 95% CI 0.02–4.63) and BA.2 (0%; 0/128, 95% CI 0.00–2.84), suggesting consistent clinical benefit with sotrovimab during the BA.2 predominant period.^32^

## Discussion

This SLR identified and assessed all observational studies in the available literature available from January 1, 2022 to February 27, 2023, which reported clinical outcomes for patients treated with sotrovimab during Omicron BA.2 and BA.5 predominance. These studies consistently reported low rates of all-cause or COVID-19-related hospitalization or death in high-risk, non-hospitalized patients receiving early treatment with sotrovimab 500 mg.

These findings build on our recently published SLR, which reviewed clinical outcomes of patients with COVID-19 treated with sotrovimab 500 mg during BA.2 subvariant predominance, and reported consistently low proportions of severe clinical outcomes (such as hospitalization and mortality) in sotrovimab-treated patients during BA.1 and BA.2 predominance.^21^ Another recent SLR and meta-analysis demonstrated the real-world effectiveness of sotrovimab for reducing hospitalization and mortality during both the Delta and Omicron BA.1 periods of predominance.^20^

Of the 14 studies included in this SLR, six high-quality studies addressed the clinical effectiveness of sotrovimab during periods of BA.2 or BA.5 predominance.^30,33,34,37–39^ Of these, two multicenter studies from the US^37^ and UK^33^ reported a lower risk of COVID-19-related hospitalization, emergency department or urgent care visits, and all-cause hospitalization or death with sotrovimab vs no treatment during BA.2 predominance in both countries. These findings support the maintained clinical effectiveness of sotrovimab against the BA.2 subvariant. In addition, although only a limited number of studies identified in our review were conducted during the period of BA.5 predominance, the findings from these four studies demonstrated low rates of COVID-19-related and all-cause clinical outcomes in sotrovimab-treated patients during this time.^27,35,36,39^ Three studies (one from Denmark and two from England) statistically compared clinical outcomes of sotrovimab-treated patients between the BA.1 and BA.2 or BA.5 predominance periods.^30,34,35^ Each found no difference in the risk of all-cause or COVID-19-related hospitalization or death during BA.2 and BA.5 predominance compared with BA.1.

Only two of the studies included in this review were conducted in the US.^26,37^ Both studies evaluated sotrovimab effectiveness during the BA.1 and BA.2 predominant periods. No data after the emergence of BA.2 were generated in the US since sotrovimab use was discontinued after April 2022 when prevalence of the BA.2 subvariant was above 50%. Consequently, all data from the BA.5 period are derived from outside the US, mainly in Europe.

Two observational cohort studies by Zheng et al. leveraged the substantial size of the OpenSAFELY platform database across BA.2 and BA.5 subvariant periods using propensity scoring methodology with sensitivity analyses to support the robustness of the data.^38,39^ In the earlier of these two studies, sotrovimab 500 mg was associated with a substantially lower risk of 28-day COVID-19-related hospitalization or death during the Omicron BA.2 subvariant surge compared with molnupiravir after adjusting for demographics, high-risk cohort categories, vaccination status, calendar time, BMI and other comorbidities (n=1970).^38^ Rates of COVID-19-related hospitalization or death for sotrovimab were comparable across the Omicron BA.1 (0.96%) and BA.2 (0.95%) periods, and mortality was lower in patients treated with sotrovimab vs molnupiravir during both periods.^38^ It should be noted, however, that between the Omicron BA.1 and BA.2 periods, guidance in the UK for molnupiravir was changed from a second-to third-line treatment option, while sotrovimab remained a first-line option during both periods.^44^ Although the effect of this change is unclear, it may have impacted the baseline characteristics of patients who received molnupiravir; the authors acknowledge the risk of bias is small.^38^

More recently, the authors reported no difference in the risk of COVID-19-related hospitalization or death between nirmatrelvir/ritonavir- and sotrovimab-treated patients during BA.2 and BA.5 predominance.^39^ The authors concluded that these data support a protective role of sotrovimab treatment against the Omicron BA.2 and BA.5 subvariants.^38,39^

The results from Zheng et al are further supported by the large retrospective cohort studies conducted by Harman et al^34^ and Patel et al.^36^ In Harman et al, variant sequencing data from patients in England were used to assess the risk of hospital admission within 14 days in patients treated with sotrovimab and infected with Omicron BA.2, compared with Omicron BA.1. Similar to Zheng et al,^38^ no significant difference in clinical outcomes was observed between BA.2 and BA.1 subvariants. The consistent results of Harman et al and Zheng et al, despite assessment of different clinical outcomes and across overlapping populations, further support the robustness of these findings. In Patel et al, consistently low COVID-19-related hospitalization rates were observed among patients receiving sotrovimab, with no evidence of significant differences in incidence rate ratio for any period compared with BA.1.^36^

There are some limitations to this study, which should be discussed. Firstly, the number of studies identified in this SLR is small, although they collectively included over 1.7 million high-risk participants. The COVID-19 landscape is also rapidly evolving and real-world data for sotrovimab during BA.2 and BA.5 predominance and onwards is still emerging. Further evidence has been published since we completed our literature search, including an OpenSAFELY population-based cohort analysis demonstrating a reduced risk of adverse outcomes among sotrovimab-treated patients versus no treatment in England during the BA.1 and BA.2 periods.^45^ In addition, a comparative effectiveness study using the DISCOVER dataset (north-west London) assessed the risk of 28-day COVID-19-related hospitalisation and/or COVID-19-related death among highest-risk patients who received sotrovimab or no early COVID-19 treatment.^46^ The risk of hospitalisation and/or death was lower for the sotrovimab-treated cohort across periods of BA.1, BA.2, and BA.5 predominance, although statistical significance was reached only for the BA.1 period. Additional observational studies will further contribute to the understanding of sotrovimab’s effectiveness during recent Omicron subvariant periods.

The observational nature of the studies included has inherent limitations, such as lack of a randomized design; however, this limitation was mitigated in many studies by use of appropriate measures to control for confounding factors. Furthermore, seven studies published in preprint databases have been included in this SLR.^33–39^ While these should be interpreted with caution as they are not peer-reviewed, preprint publication has been commonly used throughout the COVID-19 pandemic as a means of rapidly reporting outcomes in order to guide responsive public health decision-making.^47^

Due to a lack of sequencing data, most of the studies included in this SLR used an ecological design to infer the infecting variant using the date of SARS-CoV-2 infection.^26,27,30,31,33,35–39^ An exception was Mazzotta et al. and Harman et al., which used sequencing data to fully ascertain the SARS-CoV-2 subvariant of infection.^29,34^

Finally, a meta-analysis was not considered feasible as the included studies were diverse in terms of population, endpoints, study design, and analytical methods used to estimate clinical outcomes during Omicron BA.2 or BA.5. Combining studies is unwise as this may amplify the presence of confounding factors.

## Conclusions

Results from this SLR build on the findings from our earlier published review, providing further evidence for continued clinical effectiveness of early treatment with sotrovimab 500 mg IV in preventing severe clinical outcomes during Omicron BA.2 and BA.5 periods vs control/comparators and vs the Omicron BA.1 period among high-risk, non-hospitalized patients. The studies included in this review were consistent in reporting similarly low proportions of severe clinical outcomes (such as hospitalization and mortality) in sotrovimab-treated patients between the periods of Omicron BA.1, BA.2 and BA.5 subvariant predominance. Additional observational studies are warranted to contribute to the understanding of real-world effectiveness of sotrovimab against Omicron BA.2 and BA.5 subvariants, as well as future evolving variants.

## Declarations

### Author contributions

All authors took part in drafting, revising or critically reviewing the manuscript; gave final approval of the version to be published; have agreed on the journal to which the article has been submitted; and agree to be accountable for all aspects of the work in ensuring that questions related to the accuracy or integrity of any part of the work are appropriately investigated and resolved.

### Funding

This study was funded by GSK in collaboration with Vir Biotechnology, Inc (study number 220061).

### Data availability statement

All datasets generated for this study are included in this manuscript.

### Competing interests

MB is a contracted worker for GSK. EJL, DCG and MD are employees of, and/or stock/shareholders in, GSK. CR and LL are employees of PPD Evidera, which received funding from GSK and Vir Biotechnology, Inc to conduct the study.

### Ethical approval

Only publicly available papers were included in this SLR, and no human subjects were involved; ethics approval was therefore not required.

### Informed consent

Not applicable.

### Consent to participate

Not applicable.

### Consent to publish

Not applicable.

## Acknowledgments

Editorial support (in the form of writing assistance, including preparation of the draft manuscript under the direction and guidance of the authors, collating and incorporating authors’ comments for each draft, assembling tables and figures, grammatical editing, and referencing) was provided by Kathryn Wardle of Apollo, OPEN Health Communications, and was funded by GSK and Vir Biotechnology, Inc.

## References

1. World Health Organization. WHO Coronavirus (COVID-19) Dashboard. https://covid19.who.int/. Accessed 4^th^ December 2023.

2. Cucinotta D, Vanelli M. WHO Declares COVID-19 a Pandemic. Acta bio-medica : Atenei Parmensis. 2020;91(1):157–160.

3. Mendiola-Pastrana IR, López-Ortiz E, Río de la Loza-Zamora JG, González J, Gómez-García A, López-Ortiz G. SARS-CoV-2 Variants and Clinical Outcomes: A Systematic Review. *Life (Basel*, Switzerland). 2022;12(2).

4. Carabelli AM, Peacock TP, Thorne LG, et al. SARS-CoV-2 variant biology: immune escape, transmission and fitness. Nature reviews Microbiology. 2023;21(3):162–177.

5. Myers LC, Liu VX. The COVID-19 Pandemic Strikes Again and Again and Again. JAMA network open. 2022;5(3):e221760.

6. Gaudinski MR, Coates EE, Houser KV, et al. Safety and pharmacokinetics of the Fc-modified HIV-1 human monoclonal antibody VRC01LS: A Phase 1 open-label clinical trial in healthy adults. PLoS medicine. 2018;15(1):e1002493.

7. Ko SY, Pegu A, Rudicell RS, et al. Enhanced neonatal Fc receptor function improves protection against primate SHIV infection. Nature. 2014;514(7524):642–645.

8. Pinto D, Park YJ, Beltramello M, et al. Cross-neutralization of SARS-CoV-2 by a human monoclonal SARS-CoV antibody. Nature. 2020;583(7815):290–295.

9. Andrea LC, Colin H-D, Florian AL, et al. The dual function monoclonal antibodies VIR-7831 and VIR-7832 demonstrate potent in vitro and in vivo activity against SARS-CoV-2. bioRxiv. 2022:2021.2003.2009.434607.

10. Gupta A, Gonzalez-Rojas Y, Juarez E, et al. Effect of Sotrovimab on Hospitalization or Death Among High-risk Patients With Mild to Moderate COVID-19: A Randomized Clinical Trial. Jama. 2022;327(13):1236–1246.

11. GSK. GSK and Vir Biotechnology announce sotrovimab (VIR-7831) receives Emergency Use Authorization from the US FDA for treatment of mild-to-moderate COVID-19 in high-risk adults and paediatric patients. 2021; https://www.gsk.com/en-gb/media/press-releases/gsk-and-vir-biotechnology-announce-sotrovimab-vir-7831-receives-emergency-use-authorization-from-the-us-fda/#:∼:text=(Nasdaq%3A%20VIR)%20today%20announced,years%20of%20age%20and%20older. Accessed 4^th^ December 2023.

12. European Medicines Agency. Xevudy. https://www.ema.europa.eu/en/medicines/human/EPAR/xevudy. Accessed 4^th^ December 2023.

13. Australian Government: Department of Health and Aged Care. TGA provisionally approves GlaxoSmithKline’s COVID-19 treatment: sotrovimab (XEVUDY). 2021; https://www.tga.gov.au/news/media-releases/tga-provisionally-approves-glaxosmithklines-covid-19-treatment-sotrovimab-xevudy. Accessed 4^th^ December 2023.

14. Medicines & Healthcare products Regulatory Agency. Summary of Product Characteristics for Xevudy. 2022; https://www.gov.uk/government/publications/regulatory-approval-of-xevudy-sotrovimab/summary-of-product-characteristics-for-xevudy. Accessed 4^th^ December 2023.

15. Swissmedic. Swissmedic grants temporary authorisation to Xevudy® for COVID-19 patients. 2022; https://www.swissmedic.ch/swissmedic/en/home/news/coronavirus-covid-19/xevudy-fuer-covid-19-befristete-zl.html. Accessed 4^th^ December 2023.

16. World Health Organization. Weekly epidemiological update on COVID-19 - 22 March 2022. 2022; https://www.who.int/publications/m/item/weekly-epidemiological-update-on-covid-1922-march-2022. Accessed 4^th^ December 2023.

17. World Health Organization. Weekly epidemiological update on COVID-19 – 17 August 2022. 2022; https://www.who.int/publications/m/item/weekly-epidemiological-update-on-covid-1917-august-2022. Accessed 4^th^ December 2023.

18. Park YJ, Pinto D, Walls AC, et al. Imprinted antibody responses against SARS-CoV-2 Omicron sublineages. *Science (New York*, NY). 2022;378(6620):619–627.

19. US Food and Drug Administration. FDA updates Sotrovimab emergency use authorization. 2022; https://www.fda.gov/drugs/drug-safety-and-availability/fda-updates-sotrovimab-emergency-use-authorization. Accessed 4^th^ December 2023.

20. Amani B, Amani B. Efficacy and safety of sotrovimab in patients with COVID-19: A rapid review and meta-analysis. Reviews in medical virology. 2022;32(6):e2402.

21. Drysdale M, Gibbons D, Singh M, et al. Real-world effectiveness of sotrovimab for the treatment of SARS-CoV-2 infection during Omicron BA.2 subvariant predominance: a systematic literature review. medRxiv. 2023:2023.2003.2009.23287034.

22. Page MJ, McKenzie JE, Bossuyt PM, et al. The PRISMA 2020 statement: an updated guideline for reporting systematic reviews. BMJ (Clinical research ed). 2021;372:n71.

23. Higgins J, Green S. Cochrane handbook for systematic reviews of interventions. Vol 4: John Wiley & Sons; 2011.

24. Ottawa Hospital Research Institute. The Newcastle-Ottawa Scale (NOS) for assessing the quality of nonrandomised studies in meta-analyses. 2021; https://www.ohri.ca/programs/clinical_epidemiology/oxford.asp. Accessed 4^th^ December 2023.

25. Sanderson S, Tatt ID, Higgins JP. Tools for assessing quality and susceptibility to bias in observational studies in epidemiology: a systematic review and annotated bibliography. International journal of epidemiology. 2007;36(3):666–676.

26. Cheng MM, Reyes C, Satram S, et al. Real-World Effectiveness of Sotrovimab for the Early Treatment of COVID-19 During SARS-CoV-2 Delta and Omicron Waves in the USA. Infectious diseases and therapy. 2023;12(2):607–621.

27. Fujimoto K, Mutsuo S, Yasuda Y, et al. Treatment with Sotrovimab and Casirivimab/Imdevimab Enhances Serum SARS-CoV-2 S Antibody Levels in Patients Infected with the SARS-CoV-2 Delta, Omicron BA.1, and BA.5 Variants. Infectious disease reports. 2022;14(6):996–1003.

28. Martin-Blondel G, Marcelin AG, Soulié C, et al. Time to negative PCR conversion amongst high-risk patients with mild-to-moderate Omicron BA.1 and BA.2 COVID-19 treated with sotrovimab or nirmatrelvir. Clinical microbiology and infection : the official publication of the European Society of Clinical Microbiology and Infectious Diseases. 2023;29(4):543.e545–543.e549.

29. Mazzotta V, Cozzi Lepri A, Colavita F, et al. Viral load decrease in SARS-CoV-2 BA.1 and BA.2 Omicron sublineages infection after treatment with monoclonal antibodies and direct antiviral agents. Journal of medical virology. 2023;95(1):e28186.

30. Rasmussen LD, Lebech AM, Øvrehus A, et al. Experience with sotrovimab treatment of SARS-CoV-2-infected patients in Denmark. British journal of clinical pharmacology. 2023;89(6):1820–1833.

31. Zaqout A, Almaslamani MA, Chemaitelly H, et al. Effectiveness of the neutralizing antibody sotrovimab among high-risk patients with mild-to-moderate SARS-CoV-2 in Qatar. International journal of infectious diseases : IJID : official publication of the International Society for Infectious Diseases. 2022;124:96–103.

32. Nose Y, Yamamoto M. Evaluation of Safety and Clinical Outcomes of Sotrovimab in Patients Infected with SARS‒CoV‒2 in Real‒World Clinical Practice. Therapeutic Research. 2022;43(10).

33. Evans A, Qi C, Adebayo L, et al. Real-world effectiveness of molnupiravir, nirmatrelvir-ritonavir, and sotrovimab on preventing hospital admission among higher-risk patients with COVID-19 in Wales: a retrospective cohort study. medRxiv. 2023:2023.2001.2024.23284916.

34. Harman K, Nash S, Webster H, et al. Comparison of the risk of hospitalisation among BA.1 and BA.2 COVID-19 cases treated with Sotrovimab in the community in England. medRxiv. 2022:2022.2010.2021.22281171.

35. Patel V, Levick B, Boult S, et al. Characteristics and Outcomes of COVID-19 Patients Presumed to be Treated with Sotrovimab in NHS Hospitals in England. medRxiv. 2023:2023.2002.2008.23285654.

36. Patel V, Yarwood M, Levick B, et al. Characteristics and outcomes of patients with COVID-19 at high-risk of disease progression receiving sotrovimab, oral antivirals or no treatment in England. medRxiv. 2022:2022.2011.2028.22282808.

37. Young-Xu Y, Korves C, Zwain G, et al. Effectiveness of Sotrovimab in Preventing COVID-19-related Hospitalizations or Deaths Among U.S. Veterans. medRxiv. 2022:2022.2012.2030.22284063.

38. Zheng B, Green A, Tazare J, et al. Comparative effectiveness of sotrovimab and molnupiravir for prevention of severe COVID-19 outcomes in non-hospitalised patients: an observational cohort study using the OpenSAFELY platform. medRxiv. 2022:2022.2005.2022.22275417.

39. Zheng B, Tazare J, Nab L, et al. Comparative effectiveness of Paxlovid versus sotrovimab and molnupiravir for preventing severe COVID-19 outcomes in non-hospitalised patients: observational cohort study using the OpenSAFELY platform. medRxiv. 2023:2023.2001.2020.23284849.

40. Evans A, Qi C, Adebayo JO, et al. Real-world effectiveness of molnupiravir, nirmatrelvir-ritonavir, and sotrovimab on preventing hospital admission among higher-risk patients with COVID-19 in Wales: A retrospective cohort study. The Journal of infection. 2023;86(4):352–360.

41. Harman K, Nash SG, Webster HH, et al. Comparison of the risk of hospitalisation among BA.1 and BA.2 COVID-19 cases treated with sotrovimab in the community in England. Influenza and Other Respiratory Viruses. 2023;17(5):e13150.

42. Bang Z, Amelia CAG, John T, et al. Comparative effectiveness of sotrovimab and molnupiravir for prevention of severe covid-19 outcomes in patients in the community: observational cohort study with the OpenSAFELY platform. BMJ (Clinical research ed). 2022;379:e071932.

43. Ministry of Public Health – State of Qatar. Interim guidelines for management of suspected/confirmed cases of coronavirus. 2022; https://covid19.moph.gov.qa/EN/Pages/default.aspx. Accessed 4^th^ December 2023.

44. NHS England. Interim Clinical Commissioning Policy: treatments for hospital-onset COVID-19. 2022; https://www.england.nhs.uk/coronavirus/publication/interim-clinical-commissioning-policy-antivirals-or-neutralising-monoclonal-antibodies-in-the-treatment-of-hospital-onset-covid-19/. Accessed 1st September 2023.

45. The OpenSAFELY Collaborative, Tazare J, Nab L, et al. Effectiveness of sotrovimab and molnupiravir in community settings in England across the Omicron BA.1 and BA.2 sublineages: emulated target trials using the OpenSAFELY platform. medRxiv. 2023:2023.2005.2012.23289914.

46. Drysdale M, Galimov ER, Yarwood MJ, et al. Comparative effectiveness of sotrovimab versus no treatment in non-hospitalised high-risk patients with COVID-19 in North West London: a retrospective cohort study using the Discover dataset. medRxiv. 2023: 2023.07.26.23293188.

47. Watson C. Rise of the preprint: how rapid data sharing during COVID-19 has changed science forever. Nature medicine. 2022;28(1):2–5.

