## Supplemental Appendix for "Real-world effectiveness of sotrovimab for the treatment of SARS-CoV-2 infection during Omicron BA.2 and BA.5 subvariant predominance: a systematic literature review"

### SUPPLEMENTARY APPENDIX

#### Supplementary Table 1 Embase search strategy Search strategies

| **Search number** | **Search terms** |
| --- | --- |
| 1 | coronavirus disease 2019/ or (covid-19 or covid19 or corona-virus or sars-cov-2 or sars-cov2 or coronavirus disease or ncov or n-cov).ti,ab. or (alpha or beta or gamma or delta or omicron).ti. |
| 2 | sotrovimab/ or (sotrovimab or GSK-4182136 or GSK4182136 or UNII-1MTK0BPN8V or VIR-7831 or VIR7831 or Xevudy).ti,ab. |
| 3 | 1 and 2. |
| 4 | exp Clinical trial/ or exp Randomized controlled trial/ or Randomization/ or Single blind procedure/ or Double blind procedure/ or Crossover procedure/ or Placebo/ or Randomi?ed controlled trial$.tw. or Rct.tw. or Random allocation.tw. or Randomly allocated.tw. or Allocated randomly.tw. or (allocated adj2 random).tw. or Single blind$.tw. or Double blind$.tw. or ((treble or triple) adj blind$).tw. or Placebo$.tw. |
| 5 | exp longitudinal study/ or exp retrospective study/ or exp prospective study/ or exp cohort analysis/ or exp cross-sectional study/ or exp cohort analysis/ or exp observational study/ or (longitudinal study or retrospective study or prospective study or cohort$ or follow up or cross-sectional study or cross sectional study or followup study or observational study or registry or registries or real world or cross sectional or claims database or electronic health record$ or EHR or electronic medical record$ or EMR$ or RWE).ti,ab. |
| 6 | meta-analysis/ or systematic review/ or meta-analysis as topic/ or "meta analysis (topic)"/ or "systematic review (topic)"/ or exp technology assessment, biomedical/. |
| 7 | ((systematic* adj3 (review* or overview*)) or (methodologic* adj3 (review* or overview*))).ti,ab. |
| 8 | ((quantitative adj3 (review* or overview* or synthes*)) or (research adj3 (integrati* or overview*))).ti,ab. |
| 9 | ((integrative adj3 (review* or overview*)) or (collaborative adj3 (review* or overview*)) or (pool* adj3 analy*)).ti,ab. |
| 10 | (data synthes* or data extraction* or data abstraction*).ti,ab. |
| 11 | (handsearch* or hand search*).ti,ab. |
| 12 | (mantel haenszel or peto or der simonian or dersimonian or fixed effect* or latin square*).ti,ab. |
| 13 | (meta analy* or metanaly* or technology assessment* or HTA or HTAs or technology overview* or technology appraisal*).ti,ab. |
| 14 | (meta regression* or metaregression*).ti,ab. |
| 15 | (meta-analy* or metaanaly* or systematic review* or biomedical technology assessment* or bio-medical technology assessment*).mp,hw. |
| 16 | (medline or cochrane or pubmed or medlars or embase or cinahl).ti,ab,hw. |
| 17 | (cochrane or (health adj2 technology assessment) or evidence report).jw. |
| 18 | (comparative adj3 (efficacy or effectiveness)).ti,ab. |
| 19 | (outcomes research or relative effectiveness).ti,ab. |
| 20 | ((indirect or indirect treatment or mixed-treatment) adj comparison*).ti,ab. |
| 21 | or/4-20. |
| 22 | 3 and 21. |
| 23 | limit 22 to (conference abstract and yr="2022-current"). |
| 24 | limit 22 to (yr="2022-current" and (article or article in press)). |
| 25 | 23 or 24. |
